# Subcortical brain alterations in carriers of genomic copy number variants

**DOI:** 10.1101/2023.02.14.23285913

**Authors:** Kuldeep Kumar, Claudia Modenato, Clara Moreau, Christopher R. K. Ching, Annabelle Harvey, Sandra Martin-Brevet, Guillaume Huguet, Martineau Jean-Louis, Elise Douard, Charles-Olivier Martin, Nadine Younis, Petra Tamer, Anne M. Maillard, Borja Rodriguez-Herreros, Aurélie Pain, Sonia Richetin, 16p11.2 European Consortium, Simons Searchlight Consortium, Leila Kushan, Dmitry Isaev, Kathryn Alpert, Anjani Ragothaman, Jessica A. Turner, Lei Wang, Tiffany C. Ho, Lianne Schmaal, Ana I. Silva, Marianne B.M. van den Bree, David E.J. Linden, Michael J. Owen, Jeremy Hall, Sarah Lippé, Guillaume Dumas, Bogdan Draganski, Boris A. Gutman, Ida E. Sønderby, Ole A. Andreassen, Laura Schultz, Laura Almasy, David C. Glahn, Carrie E. Bearden, Paul M. Thompson, Sébastien Jacquemont

## Abstract

**Objectives:** Copy number variants (CNVs) are well-known genetic pleiotropic risk factors for multiple neurodevelopmental and psychiatric disorders (NPDs) including autism (ASD) and schizophrenia (SZ). Overall, little is known about how different CNVs conferring risk for the same condition may affect subcortical brain structures and how these alterations relate to the level of disease risk conferred by CNVs. To fill this gap, we investigated gross volume, and vertex level thickness and surface maps of subcortical structures in 11 different CNVs and 6 different NPDs.

**Methods:** Subcortical structures were characterized using harmonized ENIGMA protocols in 675 CNV carriers (at the following loci: 1q21.1, TAR, 13q12.12, 15q11.2, 16p11.2, 16p13.11, and 22q11.2) and 782 controls (Male/Female: 727/730; age-range: 6-80 years) as well as ENIGMA summary-statistics for ASD, SZ, ADHD, Obsessive-Compulsive-Disorder, Bipolar-Disorder, and Major-Depression.

**Results:** Nine of the 11 CNVs affected volume of at least one subcortical structure. The hippocampus and amygdala were affected by five CNVs. Effect sizes of CNVs on subcortical volume, thickness and local surface area were correlated with their previously reported effect sizes on cognition and risk for ASD and SZ. Shape analyses were able to identify subregional alterations that were averaged out in volume analyses. We identified a common latent dimension - characterized by opposing effects on basal ganglia and limbic structures - across CNVs and across NPDs.

**Conclusion:** Our findings demonstrate that subcortical alterations associated with CNVs show varying levels of similarities with those associated with neuropsychiatric conditions. We also observed distinct effects with some CNVs clustering with adult conditions while others clustered with ASD. This large cross-CNV and NPDs analysis provide insight into the long-standing questions of why CNVs at different genomic loci increase the risk for the same NPD, as well as why a single CNV increases the risk for a diverse set of NPDs.

## INTRODUCTION

Subcortical brain structures play a critical role in cognitive, affective, and social functions in humans (1, 2). Large-scale international neuroimaging studies have shown that major neurodevelopmental and psychiatric disorders (NPDs) (3), including schizophrenia (SZ (4)), major-depressive-disorder (MDD (5)), bipolar disorder (BD (6)), obsessive-compulsive-disorder (OCD (7)), autism-spectrum-disorder (ASD (8)), and attention-deficit-hyperactivity-disorder (ADHD (9)) are associated with alterations in subcortical structures (10–12). These case-control association studies have revealed small to moderate effect sizes on brain morphometry which have been interpreted as a consequence of heterogeneity at the level of genetics and brain mechanisms (13–16).

‘Genetics first’ studies, in which participants are ascertained based on genetic etiology, can potentially overcome challenges posed by the genetic and mechanistic heterogeneity of behaviorally defined (idiopathic) NPDs (17–19). There is a growing body of literature demonstrating subcortical volumetric alterations associated with genetic risk for NPDs as conferred by copy number variants (CNVs). CNVs are major contributors to NPDs such as ASD and SZ (15, 20), but show weaker associations with BD (21, 22) and MDD (23). Prior studies have shown that CNVs including 1q21.1-distal (24), 16p11.2-proximal BP4-5 (25), 16p11.2-distal BP2-3 (26), 15q11.2 BP1-BP2 (27), and 22q11.2 (13) affect subcortical structures with mild to large effect sizes (14). Recent studies found a significant overlap between subcortical and cortical alterations associated with 22q11.2 deletion carriers, and those associated with idiopathic schizophrenia, as well as other psychiatric illnesses (13, 28).

Beyond volumetric measurements, shape analyses of subcortical structures can capture differences that are predictive of disease status at a higher granularity (2, 29). Studies have typically focused on (13, 30): thickness defined by the distance from the medial axis of each structure, and local surface area which is a measure of surface contraction or expansion. Both shape measures have been shown to be highly heritable (31, 32), and have been used to map subcortical variation in SZ (30), ASD (29), MDD (33), and BD (34). Thickness is a proxy for subregional volume changes while the relationship between surface and volume depends on the local curvature of the region (30). For CNVs, subcortical analyses at the vertex level have only been performed in 22q11.2 deletion carriers (13) demonstrating multiple clusters of regional subcortical alterations, which were modulated by psychotic illness.

Overall, little is known about how genetic variants conferring risk for psychiatric conditions affect subcortical structures. Prior neuroimaging studies have each mainly focussed on individual CNVs, making it challenging to directly compare MRI alterations across CNVs as well as relate these MRI alterations to the level of disease risk conferred by CNVs. In particular, while Multiple CNVs confer risk for the same psychiatric conditions (35, 36), it is unknown if they are also associated with similar patterns of brain alterations underlay by a common latent dimension. Similarly, it has been shown that a common latent dimension can be identified across psychiatric diagnoses (37) and it is unknown if a similar dimension is observed for genetic risk.

Our overall aim was to systematically compare effect sizes and patterns of subcortical alterations associated with rare genetic risk for NPDs. Specifically, we aimed to: i) characterize volumetric and shape subcortical brain alterations in 11 CNVs, ii) relate effect sizes of CNVs on subcortical metrics with previously reported effects of CNVs on risk for NPDs, and iii) identify latent subcortical brain morphometry dimensions across CNVs, and NPDs.

To this end, we assembled the largest T1-weighted brain MRI dataset across all recurrent CNVs (n=11) previously associated with varying levels of risk for psychiatric illness (**Table 1**), and characterized volume, 3D surface, and thickness maps of subcortical structures. Effect sizes for 6 NPDs (ADHD, ASD, BD, MDD, OCD, and SZ) were obtained from previously published studies from the ENIGMA consortium.

**Table 1:** Demographic characteristics for all participants in the study. CNV chromosomal coordinates are provided in megabase (Mb) with the number of genes encompassed in each CNV and a well-known gene for each locus, to help recognize the CNV. Clinically ascertained participants come from five cohorts and non-clinically ascertained participants are from the UK Biobank. Diagnoses column reports the number of participants with ASD, SZ, and other diagnoses including the following list: language disorder, major depressive disorder, post traumatic stress disorder (PTSD), unspecified disruptive and impulse-control and conduct disorder, social anxiety disorder, social phobia disorder, speech sound disorder, moderate intellectual disability, specific learning disorder, gambling disorder, bipolar disorder, conduct disorder, attention deficit/hyperactivity disorder ADHD, Substance abuse disorder, global developmental delay, motor disorder, obsessive compulsive disorder, sleep disorder, Tourette’s disorder, mood disorder, eating disorders, transient tic disorder, trichotillomania, pervasive developmental disorder NOS, specific phobia, body dysmorphic disorder, mathematics disorder, dysthymic disorder. IQ loss and odds ratio (OR) for autism spectrum disorder and schizophrenia risk were extracted from previous publications (3, 51). Detailed demographics are reported in **Table ST1.** Abbreviations, Del: deletion; Dup: duplication; BC: Brain-Canada (University of Montreal); CDF: Cardiff University; EU: the 16p11.2 European Consortium; UKB: UK Biobank; SVIP: the Simons Variation in Individuals Project; ASD: autism spectrum disorder; SZ: schizophrenia; OR: Odds ratio; IQ: intelligence quotient; chr: chromosome; M: male; F: female; Age: mean age; SD: standard deviation; n Genes: number of genes encompassed by the CNV.

| Loci | Chr (hg19)<br>start-stop Mb | n Genes<br>(Gene) | Type | Ascertainment (cohorts) | N | Age<br>(SD) | Sex<br>(M/F) | Diagnoses<br>ASD/SZ/other | IQ<br>loss | OR<br>ASD/SZ |
| --- | --- | --- | --- | --- | --- | --- | --- | --- | --- | --- |
| 1q21.1 | chr1<br>146.5-147.3 | 7<br>CHD1L | Del | Clinical (EU, SVIP, BC, CDF) | 28 | 38 (21) | 22/18 | 1 / 1 / 10 | 15 | 3.2/6.4 |
|  |  |  |  | Non clinical (UKB) | 12 |  |  |  |  |  |
|  |  |  | Dup | Clinical (EU, SVIP, BC, CDF) | 17 | 47 (19) | 18/12 | 1 / 0 / 4 | 25 | 5.3/2.9 |
|  |  |  |  | Non clinical (UKB) | 13 |  |  |  |  |  |
| TAR | chr1<br>145.3-145.8 | 15<br>RBM8 | Dup | Non clinical (UKB) | 31 | 60 (8) | 14/17 | - | 2.4 | -/1 |
| 13q12.12 | chr13<br>23.5-24.8 | 5<br>SPATA13 | Dup | Non clinical (UKB) | 21 | 62 (8) | 11/10 | - | 0.6 | - |
| 15q11.2 | chr15<br>22.8-23.0 | 4<br>CYFIP1 | Del | Non clinical (UKB) | 108 | 65 (7) | 59/49 | 0 / 0 / 2 | 5.7 | 1.3/1.9 |
|  |  |  | Dup | Non clinical (UKB) | 144 | 64 (7) | 77/67 | 0 / 0 / 6 | 0.9 | 1/1 |
| 16p11.2 | chr16<br>29.6-30.2 | 27<br>KCTD13 | Del | Clinical (BC-CDF-EU-SVIP) | 78 | 19 (15) | 37/45 | 13 / 0 / 36 | 26 | 14.3/1.1 |
|  |  |  |  | Non clinical (UKB) | 4 |  |  |  |  |  |
|  |  |  | Dup | Clinical (BC-CDF-EU-SVIP) | 68 | 34 (17) | 32/43 | 10 / 1 / 19 | 11 | 10.5/11.7 |
|  |  |  |  | Non clinical (UKB) | 7 |  |  |  |  |  |
| 16p13.11 | chr16<br>15.5-16.2 | 6<br>MYH11 | Dup | Non clinical (UKB) | 50 | 66 (6) | 26/24 | - | 8.7 | 1.5/2 |
| 22q11.2 | chr22<br>19.0-21.4 | 49<br>AIFM3 | Del | Clinical (BC-CDF-UCLA) | 68 | 14 (6) | 33/35 | 8 / 2 / 31 | 28.8 | 32.3/23 |
|  |  |  | Dup | Clinical (BC-CDF-UCLA) | 19 | 29 (23) | 11/15 | 2 / 0 / 9 | 8.3 | 2/0.2 |
|  |  |  |  | Non clinical (UKB) | 7 |  |  |  |  |  |
| Controls |  |  |  | Clinical(BC-CDF-EU-SVIP-UCLA) | 317 | 48 (21) | 387/395 | 1 / 0 / 23 | - | - |
|  |  |  |  | Non clinical (UKB) | 465 |  |  |  |  |  |

## METHODS

### Participants

Recurrent deletions and duplications were included in the study if 1) the level association between the CNV and psychiatric conditions (or lack thereof) as well as the effect size on cognitive ability was previously established (15, 17, 20, 21, 38–42) and 2) MRI data were available for at least 20 carriers of the same CNV. This minimum sample size of 20 was established based on the power to detect large effect sizes as previously published (14)). Clinically ascertained groups: CNV carriers were recruited after either being referred for genetic testing due to the diagnosis of a neurodevelopmental disorder or as the relative (e.g., parent) of a CNV carrier. Controls within clinically ascertained groups were either non-carriers within the same families.

Unselected population group: CNV carriers were identified in the UK Biobank. Controls were defined as individuals who did not carry any of the 11 CNVs selected from this study.

Demographic details and coordinates of each of the 11 CNVs are provided in **Table 1**, and **Table ST1**. Signed consents were obtained by investigators from each cohort for all participants or their legal representatives prior to the investigation. This study, using an aggregate dataset, obtained ethics approval from the CHU Sainte-Justine Hospital.

### MRI image acquisition and preprocessing

The data sample included 3D T1-weighted (T1w) volumetric brain images at 0.8-1 mm isotropic resolution across all sites. MRI parameters for each cohort are detailed in the Supplemental-Material.

### Subcortical volume and shape segmentation

FreeSurfer 5.3.0 was used to segment all scans into seven bilateral subcortical regions of interest: nucleus-accumbens, amygdala, caudate, hippocampus, putamen, pallidum, and thalamus. The ENIGMA subcortical shape analysis pipeline (30) (http://enigma.ini.usc.edu/protocols/imaging-protocols/) was then applied to derive two measures of shape morphometry for each subcortical region: 1) the radial distance, which is the distance from each vertex to the medial curve of each region (referred to as thickness); 2) the logarithm of the Jacobian determinant (LogJacs), which corresponds to the surface dilation ratio between the subject structure and the template (referred to as surface). See Supplementary-Methods for details.

### Quality control

Visual quality inspection was performed by the same rater (CM) using the ENIGMA standardized quality control protocol (13).

### Normative modeling

Changes in brain measures with age (in controls; age: 6-80 years) were modeled using Gaussian processes (43), and compared with linear models (**Figure 1, Figure SF5.B**). In subsequent analyses, we used Gaussian Processes Regression (GPR, fitting a model on controls and using age, sex, site, and ICV as covariates) to obtain W-scores (GPR based Z-scores w.r.t. the mean and standard deviation modeled in controls, **Figure SF1**). See Supplementary-Methods for details.

**Figure 1:**
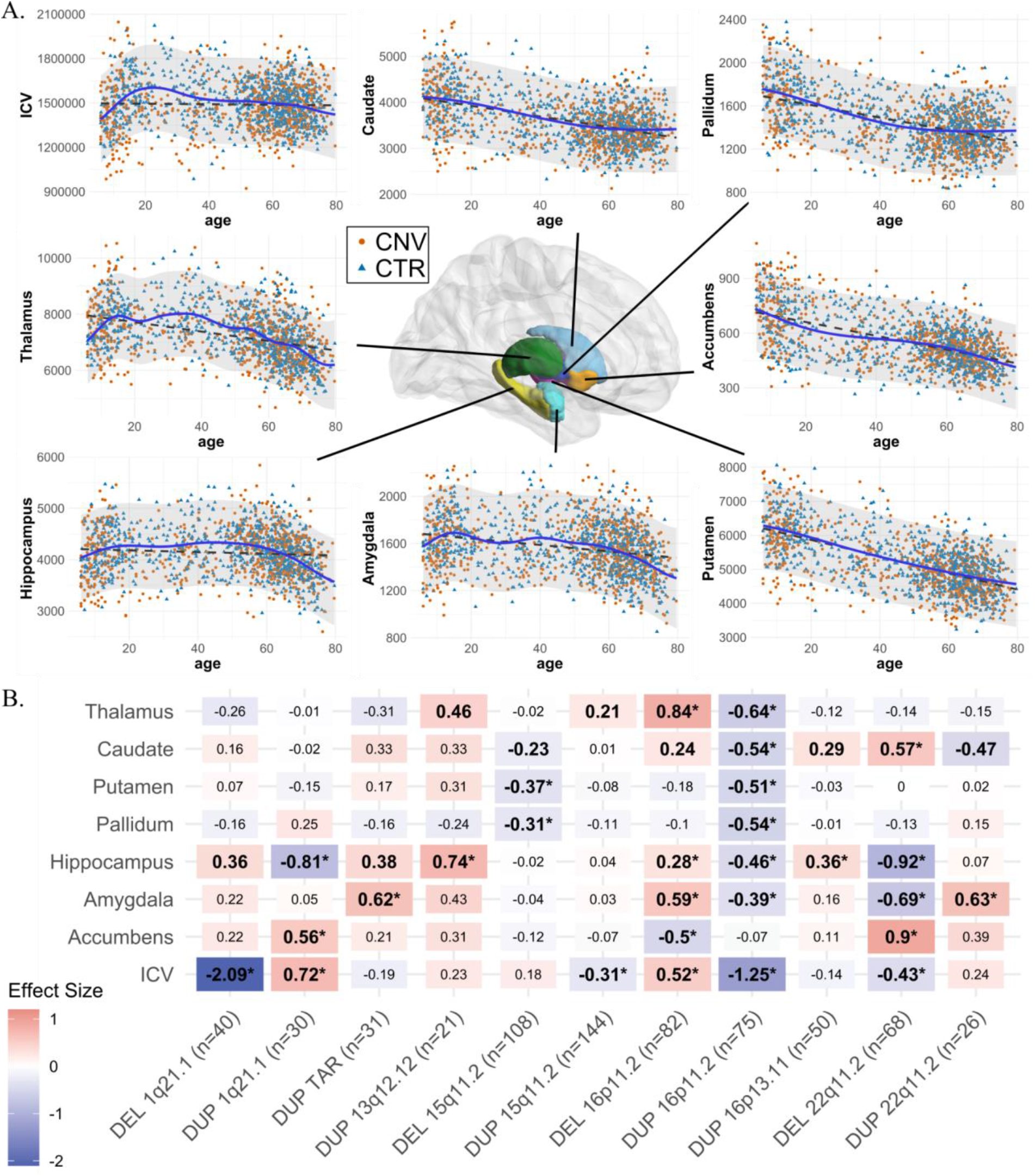
Normative age modeling and subcortical volume effect sizes. Legend: A) Scatterplots showing the distribution of ICV and subcortical volumes with age, along with Gaussian processes modeling (solid line) and a linear model (dotted line). All CNV carriers (CNV) and controls (CTR; which are used for Gaussian processes modeling) are shown as points. B) Cohen’s *d* values for subcortical structures and ICV for 11 CNVs. Case-control differences were calculated (lm function in R) using W-scores (derived from Gaussian processes modeling). W-score already includes adjustments for age, sex, site, and ICV. Significant effect sizes with nominal *p*-value <0.05 are in bold, and FDR *p*-value <0.05 are shown with an asterisk (*); FDR correction was applied across all CNVs and structures. Darker red or blue represent higher positive or negative effect sizes. Sample sizes for each analysis (for ICV) are reported in parentheses along with x-axis labels. DEL: deletions; DUP: duplications; ICV: Intracranial volume. Detailed effect sizes, standard error (SE), and *p*-values are reported in **Figure SF2**.

### Statistical analysis

Linear regression models (R version 3.6.3) were used to compute CNV-control differences (Cohen’s *d*) for each CNV using GPR based W-scores. This approach was used for CNVs across ICV, subcortical volumes, and subcortical shape analysis. The FDR procedure (44) was applied within CNVs (11-CNVs by 8-MRI volumes). For subcortical shape analysis, the FDR procedure was applied across 11-CNVs by 27000-vertices. The significance was set at FDR-corrected *q* < 0.05. See Supplementary-Methods for additional details.

### Effect sizes

Cohen’s *d* were computed based on case-control linear regression. Cohen’s *d* values for neurodevelopmental and psychiatric disorders were extracted from previous ENIGMA studies (4–9) herein referred to as ENIGMA’s Cohen’s *d* (Supplementary-Methods). All effect sizes were computed after regressing for age, sex, site, and ICV.

For comparisons across metrics, the following maximum effect sizes were used: absolute Cohen’s *d* for ICV, maximum absolute Cohen’s *d* across 7 subcortical volumes, and average absolute Cohen’s *d* of the top decile across subcortical shape vertices. Because the proportion of significant vertices varied across CNVs and NPDs (due to differences in effect and sample sizes) we chose to focus on the top decile Cohen’s *d* for all CNVs and NPDs to avoid biases and to provide effect sizes comparable across CNVs and NPDs. Vertices in the top decile were identified for thickness and surface separately and were not constrained by spatial continuity. Statistical testing of spatially correlated Cohen’s *d* profiles was performed using BrainSMASH (45, 46). See details in Supplementary-Methods.

### Quantifying shared variance across CNVs and NPDs

Principal components analysis (PCA) quantified shared variance across all CNVs and NPDs. For volume, we used CNV and NPD maps (z-scored Cohen’s *d* contrasts adjusted for ICV and nuisance variables); for vertices, we stacked the thickness and surface maps and ran a single PCA (*FactoMineR* package in R)). As a sensitivity analysis, we ran separate PCAs for CNVs and for NPDs (**Figure SF11)**. See Supplementary-Methods for additional details and methods.

## RESULTS

Effects of CNVs on subcortical volumes

Six of the 11 CNVs had significant effects on ICV. Opposing effects were observed for deletions and duplications at the same loci for 1q21.1-distal, 15q11.2, 16p11.2-proximal, and 22q11.2 (**Figure 1B**).

Nine of the 11 CNVs had significant effects on subcortical volumes. The largest effects were observed for 22q11.2 deletions followed by 16p11.2-proximal, 1q21.1-distal deletions and 1q21.1-distal, 16p11.2-proximal duplications (**Figure 1B**). Every structure was affected by at least two CNVs, and the hippocampus and amygdala were affected by five CNVs.

Sensitivity analysis testing the effect of i) the presence or absence of a psychiatric diagnosis; ii) site effects; and iii) averaging left and right subcortical volumes; demonstrated that results were robust (**Figure SF3-SF5**). In addition, the Cohen’s *d* values for 22q11.2 deletions showed high concordance (r=0.93, p=2e-3) with previously published results from a much larger overlapping sample (13) (n=68 vs n=430 deletion carriers; **Figure SF5.A**).

### Effects of CNVs on thickness and local surface area

To provide a more refined analysis of subcortical structures, we used thickness (radial distance) and surface (local-surface-area dilation/contraction). Shape analysis detected significant group differences across all CNVs, with both higher and lower thickness and local surface area relative to the control groups (**Figure 2 and Figure SF6**). The CNV-control analyses for 22q11.2 deletions and TAR duplications provided the highest and lowest number of significant vertices, respectively (**Table ST4-ST7**). For each CNV, the largest number of significant vertices was observed for thickness in caudate, and for surface in thalamus, hippocampus and caudate (**Table ST4-ST7**). A significant mirror effect at the vertex level was observed between deletions and duplication for only one locus (16p11.2, **Figure 4.A**). Because surface and thickness showed a positive correlation across most vertices in controls, we presented a simplified map representing concordant effects on surface and thickness (**Figure SF7)**. Vertices with the largest concordant increases were observed in the thalamus for 16p11.2-proximal deletions and head of the caudate for the 22q11.2 deletions. The largest concordant decreases were observed in the body and tail of the caudate, the tail of the thalamus for 16p11.2-proximal duplications, and both the head and tail of the hippocampus for 22q11.2 deletions (**Table ST8**).

**Figure 2:**
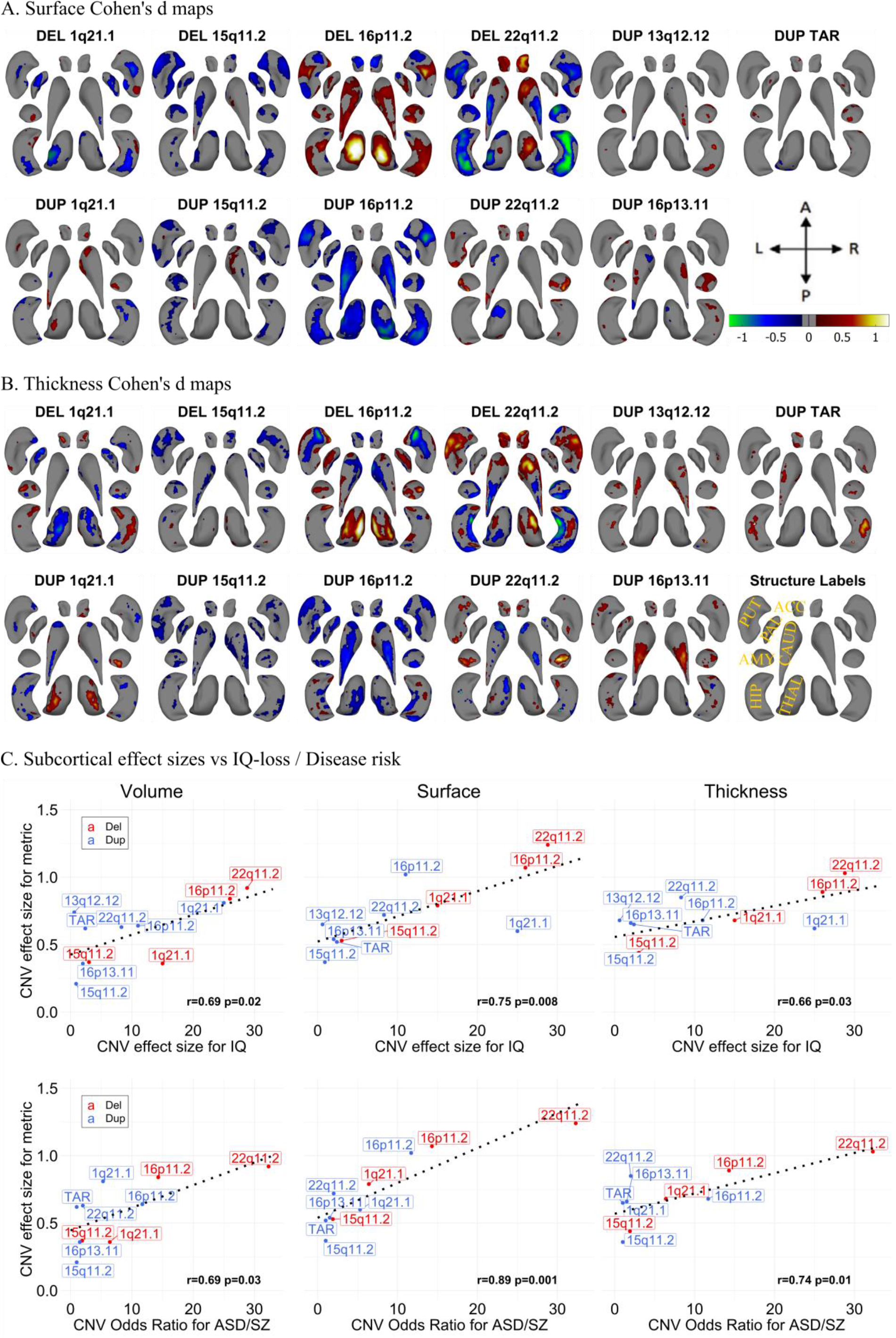
Cohen’s *d* maps for Subcortical Shape analysis and effect size comparison. Legend: A-B) Cohen’s *d* maps of subcortical shape alterations in surface (panel A); and thickness (panel B) for 11 CNVs (dorsal view). Significant vertices are shown, after applying FDR correction (<0.05) across all 27,000 vertices x 11 CNVs (within each panel). Colorbar for panels A-B are shown in panel A, and structures’ labels are shown in panel B. Thickness represents local radial distance, and surface represents local surface area dilation/contraction. Blue/green colors indicate negative coefficients, or regions with reduced thickness in the CNV group compared with the controls. Red/yellow colors indicate positive coefficients, or regions with increased thickness in the CNV group compared with the controls. Gray regions indicate areas of no significant difference after correction for multiple comparisons. Each vertex was adjusted for sex, site, age, and intracranial volume (ICV). Ventral views are shown in **Figure SF6**. Covariance as well as overlap between surface and thickness at the vertex level are shown in **Figure SF7**. C) Comparison of effect sizes of CNVs on subcortical-volume / subcortical-shape metrics and previously published effect sizes on cognition and disease risk. Regression lines fitted using the *geom_smooth* function in R. Pearson correlation and p-values (parametric *cor*.*mtest* function in R) are shown for each metric. Plots comparing the effect sizes of CNVs and the number of genes within CNV / probability of being loss-of-function intolerant (pLI-sum) for genes within CNV, as well as ICV metric are shown in **Figure SF8**. Concordance of effect sizes of CNVs on subcortical shape metrics and subcortical-volume are shown in **Figure SF9.** Abbreviations, DEL: deletion; DUP: duplication; ACC: accumbens; AMY: amygdala; CAUD: caudate; HIP: hippocampus; PUT: putamen; PAL: pallidum; THAL: thalamus; ES: effect size; CCC: concordance correlation coefficients; Directions: L-left, R-right, A-anterior, P-posterior.

Effect sizes of CNVs on thickness and surface were concordant with those reported for volume (CCC=0.68, p=0.006; and CCC=0.57, p=0.02 respectively), but were on average higher (bias-factor = 0.88 and 0.85, respectively) (**Figure SF9**, and **Table ST3**).

### CNV effect sizes on subcortical volume/shape, cognition and risk for disease

We showed that the effect sizes of CNVs on subcortical volume, thickness and surface were 2-to 6-fold larger than those previously published in the ENIGMA studies of idiopathic ADHD, ASD, BD, MDD, OCD, and SZ (volume) and MDD, SZ (for shape metrics) (e.g., the largest effects for 22q11.2 deletion/SZ were 0.92/0.46 for volume, 1.03/0.39 for thickness; and 1.24/0.34 for surface respectively; **Table ST2**).

We then asked if CNV effect sizes were related to their effects on cognition and disease risk. We observed a significant correlation between the effect size of CNVs on subcortical volumes/thickness/surface and their previously reported effect size on IQ (3, 38, 47) (r=0.66-0.75, p<0.03) as well as risk for either ASD (3, 20, 48, 49) or SZ (3, 15, 49) (r=0.69-0.89, p<0.03, **Figure 2C**). Effect size on subcortical structure was also correlated to the gene content (measured by pLI or number of genes) of each CNV (**Figure SF8**). On the other hand, CNV effect sizes on ICV were not significantly associated with cognition (**Figure SF6**).

### Comparing Cohen’s *d* profiles of CNVs and NPDs

Because CNVs are pleiotropic, conferring risk for multiple neuropsychiatric conditions (50), we investigated whether there were any similarities between profiles of subcortical alterations across CNVs and NPDs. We correlated subcortical volume effect sizes across CNVs and NPDs, and performed a hierarchical clustering analysis (using Ward’s method). Schizophrenia was part of a cluster including BD, MDD, OCD, 22q11.2 deletions and 1q21.1-distal duplications. This cluster was negatively correlated with the cluster encompassing ASD, 16p11.2-proximal deletions, 15q11.2 deletions, 15q11.2 duplications, and 13q12.12 duplications. ADHD did not cluster with any of the conditions or CNVs (**Figure 3A**).

**Figure 3:**
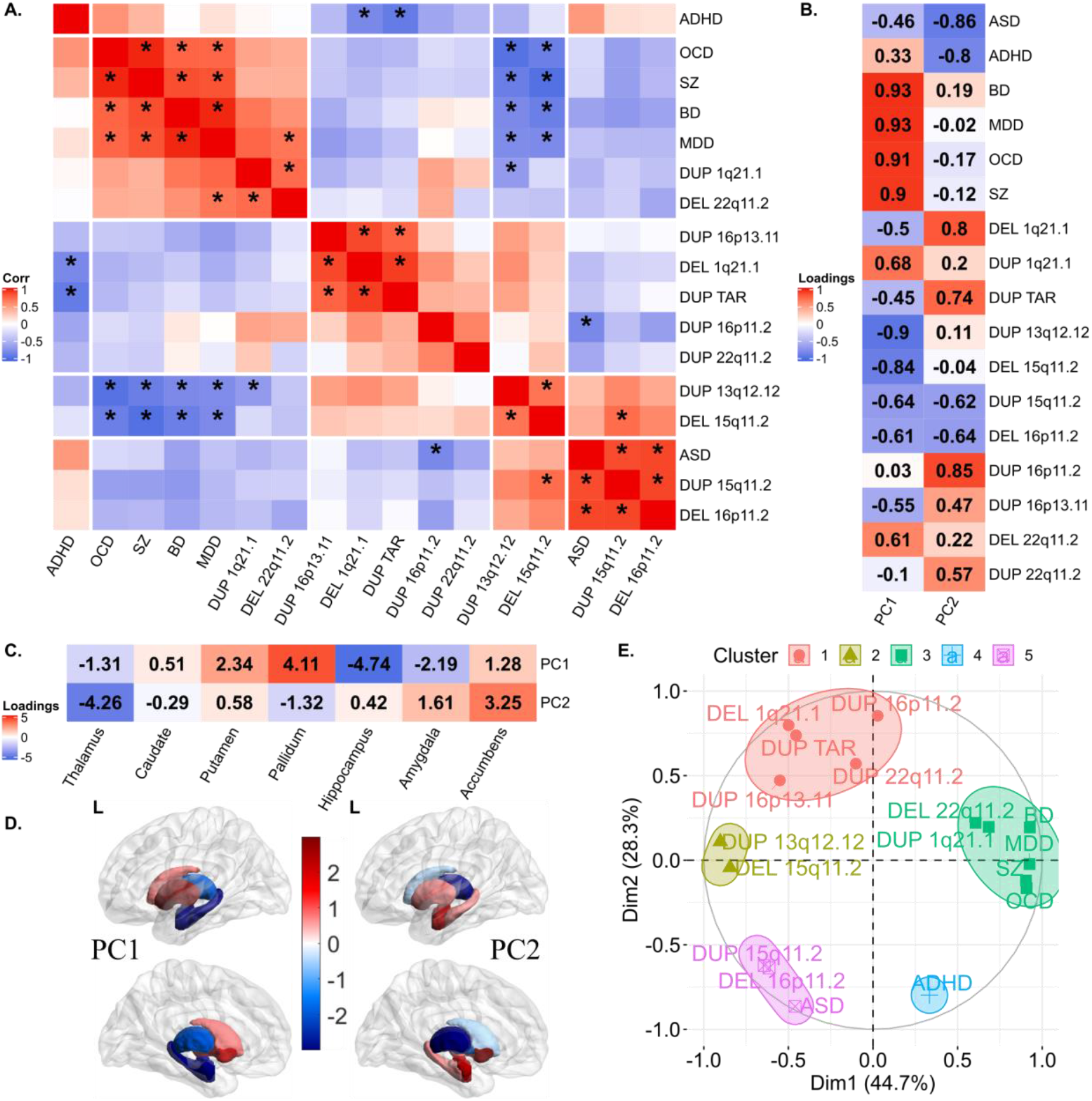
Correlations and principal components analysis across CNVs and NPDs. Legend: A) Correlations between Cohen’s *d* profiles of CNVs and NPDs. * represent *p*-value <0.05 (BrainSMASH). Hierarchical (Ward distance) clustering based 5 clusters are separated using white spaces. B-E) Principal components analysis across subcortical volumes of 11 CNVs and 6 NPDs. B) Variable loadings on PC1 and PC2; C) Subcortical structures’ loadings; D) PC1 and PC2 loadings mapped on subcortical structures. (E) Correlation circle showing CNVs and NPDs in PC1 and PC2 space. CNV-NPD groupings obtained using *K*-means clustering (k=5 clusters) in the PC space (Euclidean distance). Abbreviations, CNV: copy number variants; DEL: deletion; DUP: duplication; NPD: neurodevelopmental and psychiatric disorders; Corr: Pearson correlation; ASD: autism spectrum disorder; ADHD: attention deficit hyperactivity disorder; BD: bipolar disorder; MDD: major depressive disorder; OCD: obsessive-compulsive disorder; SZ: schizophrenia; PC: principal component; L: left hemisphere; Dim: dimension.

However, there was no clear relationship between the level of ASD or SZ risk conferred by CNVs and their clustering with those 2 conditions (correlation between CNVs risk for ASD and CNVs-ASD clustering r=-0.14, (p=0.7); correlation between CNVs risk for SZ and CNVs-SZ clustering: r=0.59, (p=0.07)) (**Figure SF10**).

### Subcortical latent dimensions across CNVs and NPDs

To investigate the clusters observed above, we performed a PCA on profiles of subcortical volume effect sizes. The first two principal components (PCs) explained 45% and 28% of the variance in Cohen’s *d* values. Dimension 1 of the NPDs and CNVs showed positive and negative loadings for the basal ganglia (pallidum, putamen) and limbic system (thalamus, hippocampus, amygdala), respectively (**Figure 3C**). The second PC dimension was characterized by the accumbens and thalamus loading on both extremes. Five clusters were obtained by running K-means clustering using PC1 and PC2, with cluster 3 (green-color) corresponding to adult NPDs, cluster 4 and 5 corresponding to ADHD, and ASD respectively, and cluster 1 and 2 to CNVs. These groupings were reflected in the correlation matrix of Cohen’s *d* profiles (**Figure 3A**). To test if CNVs and NPDs separately resulted in similar dimensions, we performed independent PCAs on NPDs and CNVs. Latent dimensions (PCs) of both independent PCAs were highly correlated with each other (r= -0.93 to -0.83) (**Figure SF11**).

### Latent dimensions across subcortical shape metrics of CNVs and NPDs

To understand potentially shared and distinct effects across CNVs, we performed a multivariate analysis (PCA) on Cohen’s *d* maps of both subcortical thickness, and local-surface-area for 11 CNVs, and ENIGMA maps for SZ and MDD (**Figure 4**, and **Figure SF12**).

PCs identified positive and negative loadings for regions within the same structures. Ventral and dorsal regions showed distinct patterns of alterations. Alterations were mostly bilateral except for the thalamus. As an example, for PC1, vertices with concordant effects (same directionality for thickness and surface) suggested a decrease in subregional volumes of the body and an increase in the head and tail of the caudate. For the hippocampus, most vertices with concordant alterations suggested a volume decrease in the body, but focal increases were also observed in the head (PC1) and tail (PC2), (**Figure 4E-H**, and **Table ST9**).

**Figure 4:**
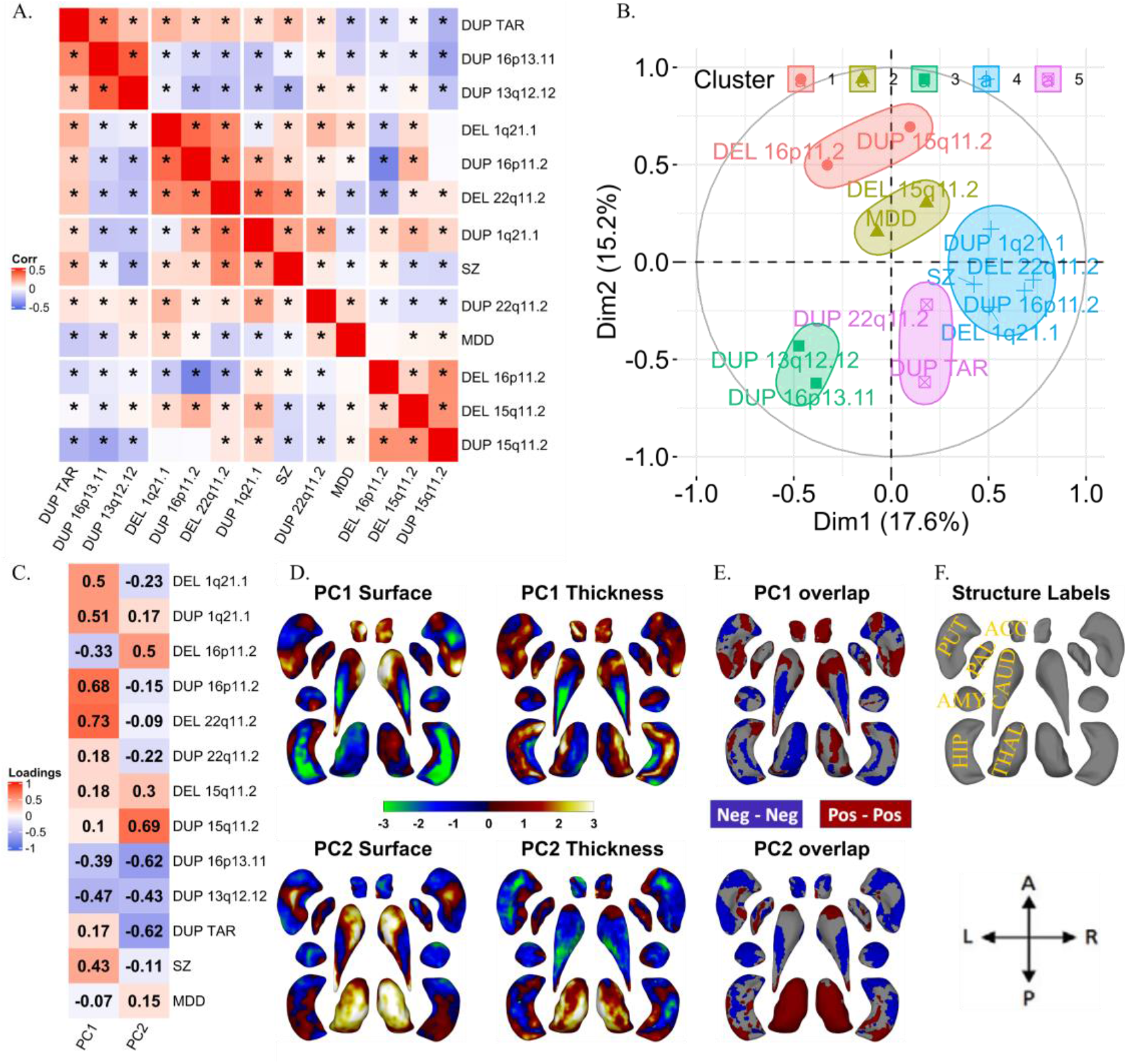
Correlations and principal components analysis across vertex-wise Cohen’s d maps of CNVs and NPDs. Legend: A) Correlations between vertex-wise Cohen’s d profiles of CNVs and NPDs. * represent p-value <0.01 (parametric test). Hierarchical (Ward distance) clustering based 5 clusters are separated using white spaces. B) Correlation circles with CNV and NPD clusters in PC1-PC2 space; C) CNV and NPD loadings of principal components 1 and 2.; D) PC1 and PC2 brain maps (dorsal views); E) overlap of PCs of thickness and surface; F) structures’ labels and dorsal view directions. Thickness represents local radial distance, and surface represents local surface area dilation/contraction. Principal components analysis was run with CNVs as variables and vertices as observations (stacked across surface and thickness metric and all subcortical structures; Z-scored). For PC maps, blue/green and red/yellow colors indicate negative and positive coefficients respectively. For overlap maps, blue and red represent negative-negative / positive-positive thickness and surface PC loadings at each vertex respectively. Ventral views are shown in **Figure SF12.** Abbreviations, DEL: deletion; DUP: duplication; PC: principal component; Dim: dimension; MDD: major depressive disorder; SZ: schizophrenia; ACC: accumbens; AMY: amygdala; CAUD: caudate; HIP: hippocampus; PUT: putamen; PAL: pallidum; THAL: thalamus; Directions: L-left, R-right, A-anterior, P-posterior.

Comparing PCA results across subcortical metrics shows that structures contributing to the latent dimension for volume are on average also those contributing to latent dimension for surface and thickness (**Figure SF13**). Lower variance explained for shape analyses suggests that shared Cohen’s d profiles may decrease when moving from gross volume to measures with higher granularity (**Figure SF14-15**). To formally test this hypothesis, we randomly sampled a smaller number of vertices and re-ran the PCA. The resulting analysis shows that variance explained for surface+thickness by two principal components decreases from 52% (for n=10 vertices), to 32% (n=1000 vertices) and stabilized from n=1000 to n=2×27200 vertices for surface+thickness (**Figure SF16**).

## DISCUSSION

This large neuroimaging study characterizing and comparing the subcortical alteration associated with 11 CNVs and six NPDs detects effects on subcortical volumes in nine out of 11 CNVs. Analyses at a higher granularity using shape metrics showed that these effects were localized to subregions of the subcortical structures. The effect sizes of CNVs on subcortical structures were correlated with their previously reported effect size on cognition and risk for ASD and SZ. That is, larger and gene-rich CNVs (e.g., 22q11.2 and 16p11.2-proximal) - which show higher risk for disease - also showed greater alterations of subcortical structures. Cohen’s *d* for CNVs were larger than those derived from case-control association studies for group of individuals with a psychiatric condition. In line with the pleiotropic effects of CNVs on risk for multiple NPDs, we identified latent dimensions explaining 44.7% of the variance in Cohen’s *d* maps across all CNVs and NPDs. Latent dimensions were defined by opposing loadings on basal ganglia and limbic structures.

All 11 CNVs showed significant effects on subcortical thickness and local surface area and most of them also affected subcortical volume. The largest effects across metrics were observed for 22q11.2 deletions. Hippocampus volume was altered by 5 CNVs, and had the largest number of significant vertices for surface across CNVs. The effect sizes of CNVs on volume, surface and thickness were correlated with the previously reported mean effect size of each CNV on cognition and risk for disease. The same correlation has been reported between effect sizes of CNVs on functional connectivity metrics and cognition / risk for diseases (51). However, there was no relationship between effect sizes on ICV and risk for disease or cognition (14) indicating that the increase in granularity achieved by vertex level shape analysis could improve our understanding of brain-behavior relationships. Studying brain-behavior relationships in unselected populations has proven to be very challenging with very small effect sizes (52). As opposed to fitting an average MRI pattern across a cognitive dimension, our alternative genetic first approach provides a much stronger correlation by working with multiple MRI-profiles associated with each genetic variant and their respective behavioral alterations. In other words, this allows one to move from single to multiple modes of brain-behavior associations.

Most of the variance in Cohen’s *d* profiles for subcortical structures was distinct for each CNV. This is consistent with a recent study showing relative specificity of association between CNVs and brain alterations using morphometric similarity mapping of cortical regions (53). On the other hand, the amount of variance explained by PCs (PC1= 45% and PC2=28%) is similar to those (32% and 29%) previously published for cortical regions-of-interest (ROI), cortical thickness and surface area for eight CNVs (54). This presence of shared effects across CNVs, regardless of brain measures analyzed, suggests that similar brain mechanisms might underlie brain alterations across subsets of CNVs.

Extending the principal components analysis (PCA) across CNVs and NPDs identified components similar to the ones described above, defined by opposing loadings on basal ganglia and limbic structures. Basal ganglia and limbic structures were previously identified as structures delineating different SZ subtypes using data driven approaches (34). These shared Cohen’s *d* profiles may explain some of the pleiotropic effects of CNVs (i.e. why all NPD CNVs increase risk for either ASD or SZ, or for both conditions). Subcortical structures with top PC loading include basal ganglia structures known to be primarily involved in motor control (55), as well as limbic structures involved in higher order functions including motivation, emotion, learning, and memory, reflecting a unimodal to transmodal axis (56). The principal component identified across subcortical structures for CNVs and NPDs, should be further investigated in relationship with the sensorimotor-association axis reported for cortical organization (56, 57).

Our findings also suggest significant heterogeneity across CNVs and NPDs. CNVs showed either little or even negative correlations with NPDs. The latter tend to cluster among themselves, except for ASD and ADHD (37). For example, 16p11.2 duplications -which increase risk for SZ and ASD-showed effects on subcortical shapes that were negatively correlated with those observed for idiopathic ASD. This could suggest that within a group of individuals with the same psychiatric diagnosis, some may show opposing MRI alterations. In addition, while effect size of CNVs on subcortical structures were correlated to risk, there was no concordance between the level of ASD- or SZ-risk conferred by CNVs and the similarity between their subcortical profiles. This highlights again, multiple modes of brain-behavior association underlying the heterogeneity of brain patterns associated with a psychiatric condition.

While some structures showed no significant differences at the gross volume level, shape analyses of the same structures revealed regions of lower as well as greater surface and thickness. For example, while the volume of the caudate has small loading on PCs, sub-regions of the caudate were top contributors to PCs of shape metrics. This finding highlights the relevance of analyzing subcortical structures at the vertex level, which may identify alterations that are averaged out at the gross volumetric level.

Sub-regions of the hippocampus that were among the top contributors to the PCA performed across CNVs– were also previously reported in shape analysis of MDD (33) and SZ (30). The observed shape differences have been reported to reflect patterns of neuronal deficits in postmortem studies of individuals with schizophrenia (30). For example, the mixed findings for caudate - increase and decrease for shape metrics in sub-regions - reflect findings from postmortem studies showing both larger (58) as well as smaller disease-related changes in total neuron number (59). The PC1 loadings of gross volume correlated with mean PC loadings for shape analysis, reflecting a consistent latent dimension. The microscopic reductions in neuron size or total number of neurons, that reflect shape differences, may manifest in macroscopic reductions in volume measured by MRI (30).

While shared variation could have been influenced by clinical ascertainment or psychiatric diagnoses, sensitivity analyses showed that this is not the case, consistent with previous publications (54). Larger sample sizes with complete balanced coverage of the lifespan age range will be required for more accurate normative modeling (60). Direct comparison of shape metrics between CNVs and psychiatric conditions is required to identify latent dimensions across CNVs and conditions and will be the focus of future studies.

In summary, effect sizes of CNVs on subcortical structures were correlated with their effect size on cognition and risk for disease. Shape analyses highlighted subregional volume alterations that were averaged out in global volume analyses. Principal components captured common effects on subcortical volumes across CNVs and NPDs may underly some of the pleiotropic effects of CNVs. Basal ganglia and limbic structures contributed to the latent dimension for volume are also those contributing to latent dimension for surface and thickness.

## Data Availability

UK Biobank data was downloaded under the application 40980, and may be accessed via their standard data access procedure (see http://www.ukbiobank.ac.uk/register-apply). UK Biobank CNVs were called using the pipeline developed in the Jacquemont Lab, as described at https://github.com/labjacquemont/MIND-GENESPARALLELCNV. The final CNV calls are available for download from the UK Biobank returned datasets (Return ID: 3104, https://biobank.ndph.ox.ac.uk/ukb/dset.cgi?id=3104). References to the processing pipeline and R package versions used for analysis are listed in methods.

## Author contributions

C.Mod., K.K., C.C, P.M.T., C.E.B., and S.J. designed the study, analyzed imaging data, and drafted the manuscript.

### Analyses

C.Mod. and C.C. performed all the preprocessing. K.K. and C.Mod. performed all the analyses of neuroimaging data. G.D. contributed to normative modeling. B.G. contributed to shape analysis. C.C, C.Mor., P.M.T., and C.E.B. contributed to result interpretation and in the editing of the manuscript.

### Data collection

C.Mod., A.M., B.R-H., A.P., S.R., and S.M-B. recruited and scanned participants in the 16p11.2 European Consortium. S.L., C.O.M., N.Y., P.T., E.D., F. T-D., V.C., A.R.C., F.D. recruited and scanned participants in the Brain Canada cohort. L.K., C.E.B collected and provided the data for the UCLA cohort. D.E.J.L., M.J.O., M.B.M. V.d.B., J.H. and A.I.S., provided the data for the Cardiff cohort.

All authors provided feedback on the manuscript.

## Data and cohort Information

Each cohort and corresponding study received approval from their local institutional review board and this study was approved by the Institutional review board of the CHU Ste Justine research center. The Simons Searchlight Consortium principal investigator is Wendy K. Chung. Contributors to the Simons Searchlight Consortium include the following: Hanalore Alupay, BS, Benjamin Aaronson, BS, Sean Ackerman, MD, Katy Ankenman, MSW, Ayesha Anwar, BA, Constance Atwell, PhD, Alexandra Bowe, BA, Arthur L. Beaudet, MD, Marta Benedetti, PhD, Jessica Berg, MS, Jeffrey Berman, PhD, Leandra N. Berry, PhD, Audrey L. Bibb, MS, Lisa Blaskey, PhD, Jonathan Brennan, PhD, Christie M. Brewton, BS, Randy Buckner, PhD, Polina Bukshpun, BA, Jordan Burko, BA, Phil Cali, EdS, Bettina Cerban, BA, Yishin Chang, MS, Maxwell Cheong, BE, MS, Vivian Chow, BA, Zili Chu, PhD, Darina Chudnovskaya, BS, Lauren Cornew, PhD, Corby Dale, PhD, John Dell, BS, Allison G. Dempsey, PhD, Trent Deschamps, BS, Rachel Earl, BA, James Edgar, PhD, Jenna Elgin, BS, Jennifer Endre Olson, PsyD, Yolanda L Evans, MA, Anne Findlay, MA, Gerald D Fischbach, MD, Charlie Fisk, BS, Brieana Fregeau, BA, Bill Gaetz, PhD, Leah Gaetz, MSW, BSW, BA, Silvia Garza, BA, Jennifer Gerdts, PhD, Orit Glenn, MD, Sarah E Gobuty, MS, CGC, Rachel Golembski, BS, Marion Greenup, MPH, MEd, Kory Heiken, BA, Katherine Hines, BA, Leighton Hinkley, PhD, Frank I. Jackson, BS, Julian Jenkins III, PhD, Rita J. Jeremy, PhD, Kelly Johnson, PhD, Stephen M. Kanne, PhD, Sudha Kessler, MD, Sarah Y. Khan, BA, Matthew Ku, BS, Emily Kuschner, PhD, Anna L. Laakman, MEd, Peter Lam, BS, Morgan W. Lasala, BA, Hana Lee, MPH, Kevin LaGuerre, MS, Susan Levy, MD, Alyss Lian Cavanagh, MA, Ashlie V. Llorens, BS, Katherine Loftus Campe, MEd, Tracy L. Luks, PhD, Elysa J. Marco, MD, Stephen Martin, BS, Alastair J. Martin, PhD, Gabriela Marzano, HS, Christina Masson, BFA, Kathleen E. McGovern, BS, Rebecca McNally Keehn, PhD, David T. Miller, MD, PhD, Fiona K. Miller, PhD, Timothy J. Moss, MD, PhD, Rebecca Murray, BA, Srikantan S. Nagarajan, PhD, Kerri P. Nowell, MA, Julia Owen, PhD, Andrea M. Paal, MS, Alan Packer, PhD, Patricia Z. Page, MS, Brianna M. Paul, PhD, Alana Peters, BS, Danica Peterson, MPH, Annapurna Poduri, PhD, Nicholas J. Pojman, BS, Ken Porche, MS, Monica B. Proud, MD, Saba Qasmieh, BA, Melissa B. Ramocki, MD, PhD, Beau Reilly, PhD, Timothy P. L. Roberts, PhD, Dennis Shaw, MD, Tuhin Sinha, PhD, Bethanny Smith-Packard, MS, CGC, Anne Snow Gallagher, PhD, Vivek Swarnakar, PhD, Tony Thieu, BA, MS, Christina Triantafallou, PhD, Roger Vaughan, PhD, Mari Wakahiro, MSW, Arianne Wallace, PhD, Tracey Ward, BS, Julia Wenegrat, MA, and Anne Wolken, BS. European 16p11.2 Consortium principal investigator Sébastien Jacquemont. Members of the European 16p11.2 Consortium include the following: Addor Marie-Claude, Service de génétique médicale, Centre Hospitalier Universitaire Vaudois, Lausanne University, Switzerland; Andrieux Joris, Institut de Génétique Médicale, CHRU de Lille, Hopital Jeanne de Flandre, France; Arveiler Benoît, Service de génétique médicale, CHU de Bordeaux-GH Pellegrin, France; Baujat Geneviève, Service de Génétique Médicale, CHU Paris - Hôpital Necker-Enfants Malades, France; Sloan-Béna Frédérique, Service de médecine génétique, Hôpitaux Universitaires de Genève - HUG, Switzerland; Belfiore Marco, Service de génétique médicale, Centre Hospitalier Universitaire Vaudois, Lausanne University, Switzerland; Bonneau Dominique, Service de génétique médicale, CHU d’Angers, France; Bouquillon Sonia, Institut de Génétique Médicale, Hopital Jeanne de Flandre, Lille, France; Boute Odile, Hôpital Jeanne de Flandre, CHRU de Lille, Lille, France; Brusco Alfredo, Genetica Medica, Dipartimento di Scienze Mediche, Università di Torino, Italy; Busa Tiffany, Département de génétique médicale, CHU de Marseille, Hôpital de la Timone, France; Caberg Jean-Hubert, Centre de génétique humaine, CHU de Liège, Belgique; Campion Dominique, Service de psychiatrie, Centre hospitalier de Rouvray, Sotteville lès Rouen, France; Colombert Vanessa, Service de génétique médicale, Centre Hospitalier Bretagne Atlantique CH Chubert-Vannes, France; Cordier Marie-Pierre, Service de génétique clinique, CHU de Lyon, Hospices Civils de Lyon, France; David Albert, Service de Génétique Médicale, CHU de Nantes, Hôtel Dieu, France; Debray François-Guillaume, Service de Génétique Humaine, CHU Sart Tilman - Liège, Belgique; Delrue Marie-Ange, Service de génétique médicale, CHU de Bordeaux, Hôpital Pellegrin, France; Doco-Fenzy Martine, Service de Génétique et Biologie de la Reproduction, CHU de Reims, Hôpital Maison Blanche, France; Dunkhase-Heinl Ulrike, Department of Pediatrics, Aabenraa Hospital, Sonderjylland, Denmark; Edery Patrick, Service de génétique clinique, CHU de Lyon, Hospices Civils de Lyon, France; Fagerberg Christina, Department of Clinical Genetics, Odense University hospital, Denmark; Faivre Laurence, Centre de génétique, Hôpital d’Enfants, CHU Dijon Bourgogne - Hôpital François Mitterrand, France; Forzano Francesca, Ambulatorio di Genetica Medica, Ospedali Galliera di Genova, Italy and Clinical Genetics Department, 7^th^ Floor Borough Wing, Guy’s Hospital, Guy’s & St Thomas’ NHS Foundation Trust, Great Maze Pond, London SE1 9RT, UK; Genevieve David, Département de Génétique Médicale, Maladies Rares et Médecine Personnalisée, service de génétique clinique, Université Montpellier, Unité Inserm U1183, CHU Montpellier, Montpellier, France; Gérard Marion, Service de Génétique, CHU de Caen, Hôpital Clémenceau, France; Giachino Daniela, Genetica Medica, Dipartimento di Scienze Cliniche e Biologiche, Università di Torino, Italy; Guichet Agnès, Service de génétique, CHU d’Angers, France; Guillin Olivier, Service de psychiatrie, Centre hospitalier du Rouvray, Sotteville lès Rouen, France; Héron Delphine, Service de Génétique clinique, CHU Paris-GH La Pitié Salpêtrière-Charles Foix - Hôpital Pitié Salpêtrière, France; Isidor Bertrand, Service de Génétique Médicale, CHU de Nantes, Hôtel Dieu, France; Jacquette Aurélia, Service de Génétique clinique, CHU Paris-GH La Pitié Salpêtrière-Charles Foix - Hôpital Pitié-Salpêtrière, France; Jaillard Sylvie, Service de Génétique Moléculaire et Génomique – Pôle biologie, CHU de Rennes, Hôpital Pontchaillou, France; Journel Hubert, Service de génétique médicale, Centre Hospitalier Bretagne Atlantique CH Chubert-Vannes, France; Keren Boris, Centre de Génétique Moléculaire et Chromosomique, CHU Paris-GH La Pitié Salpêtrière-Charles Foix - Hôpital Pitié-Salpêtrière, France; Lacombe Didier, Service de génétique médicale, CHU de Bordeaux-GH Pellegrin, France; Lebon Sébastien, Pediatric Neurology Unit, Department of Pediatrics, Lausanne University Hospital, Lausanne, Switzerland; Le Caignec Cédric, Service de Génétique Médicale - Institut de Biologie, CHU de Nantes, France; Lemaître Marie-Pierre, Service de Neuropédiatrie, Centre Hospitalier Régional Universitaire de Lille, France; Lespinasse James, Service génétique médicale et oncogénétique, Hotel Dieu, Chambéry, France; Mathieu-Dramart Michèle, Service de Génétique Clinique, CHU Amiens Picardie, France; Mercier Sandra, Service de Génétique Médicale, CHU de Nantes, Hôtel Dieu, France; Mignot Cyril, Service de Génétique clinique, CHU Paris-GH La Pitié Salpêtrière-Charles Foix - Hôpital Pitié-Salpêtrière, France; Missirian Chantal, Département de génétique médicale, CHU de Marseille, Hôpital de la Timone, France; Petit Florence, Service de génétique clinique Guy Fontaine, Hôpital Jeanne de Flandre, CHRU de Lille, France; Pilekær Sørensen Kristina, Department of Clinical Genetics, Odense University Hospital, Denmark; Pinson Lucile, Département de Génétique Médicale, Maladies Rares et Médecine Personnalisée, service de génétique clinique, Université Montpellier, Unité Inserm U1183, CHU Montpellier, Montpellier, France; Plessis Ghislaine, Service de Génétique, CHU de Caen, Hôpital Clémenceau, France; Prieur Fabienne, Service de génétique clinique, CHU de Saint-Etienne - Hôpital Nord, France; Raymond Alexandre, Center for Integrative Genomics, Lausanne University, Switzerland; Rooryck-Thambo Caroline, Laboratoire de génétique moléculaire, CHU de Bordeaux-GH Pellegrin, France; Rossi Massimiliano, Service de génétique clinique, CHU de Lyon, Hospices Civils de Lyon, France; Sanlaville Damien, Laboratoire de Cytogénétique Constitutionnelle, CHU de Lyon, Hospices Civils de Lyon, France; Schlott Kristiansen Britta, Department of Clinical Genetics, Odense University Hospital, Denmark; Schluth-Bolard Caroline, Laboratoire de Cytogénétique Constitutionnelle, CHU de Lyon, Hospices Civils de Lyon, France; Till Marianne, Service de génétique clinique, CHU de Lyon, Hospices Civils de Lyon, France; Van Haelst Mieke, Department of Genetics, University Medical Center Utrecht, Holland; Van Maldergem Lionel, Centre de Génétique humaine, CHRU de Besançon - Hôpital Saint-Jacques, France.

