## Supplement for "Subcortical brain alterations in carriers of genomic copy number variants"

### Supplement: Subcortical brain alterations in CNVs

INDEX

**MRI data acquisition and quality check** 3
Participants 3
16p11.2 European Consortium 3
Simons Searchlight Consortium 4
Brain Canada (BC, University of Montreal) 4
UCLA 5
Cardiff 5
MRI quality control 5
List of abbreviations 6

**Supplementary Methods** 6
Subcortical volume and shape segmentation 6
Subcortical volume and shape Cohen's d maps from ENIGMA: 7
Rationale for computing maximum effect sizes for subcortical shape metrics 7
Normative modeling 7
Site impact on Normative modeling 8
Statistical testing of Cohen's d profiles (BrainSMASH) 8
Covariance between shape metrics: Inter-individual maps of variation 8
Significance of the variance explained in Principal Component Analysis 8
Impact of the number of vertices on variance explained 9

**Supplementary Tables** 10
Supplement Table 1: Detailed demographics 10
Supplement Table 2: Summary of effect sizes across CNVs and NPDs, for MRI-metrics. 11
Supplement Table 3: Concordance between CNV effect size across metrics: ICV-volume-surface-thickness. 12
Supplement Table 4: Number of significant vertices per ROI across 11 CNVs for thickness. 13
Supplement Table 5: Mean of absolute Cohen's d for significant vertices per ROI across 11 CNVs for thickness. 13
Supplement Table 6: Number of significant vertices per ROI across 11 CNVs for local surface area. 14
Supplement Table 7: Mean of absolute Cohen's d for significant vertices per ROI across 11 CNVs for local surface area. 15
Supplement Table 8: Number of surface and thickness overlap vertices per ROI across 11 CNVs. 15
Supplement Table 9: Number of surface and thickness PC overlap vertices per ROI. 16

**Supplementary Figures** 17
Supplement Figure 1: W-score obtained using Gaussian Processes modeling. 17
Supplement Figure 2: Effect sizes, SE, and p-values for subcortical volumes and ICV. 18
Supplement Figure 3: Sensitivity analysis- bilateral effect sizes (Cohen's d) for subcortical structures and ICV. 19
Supplement Figure 4: Impact of the site on Gaussian Processes modeling (leave one site out analysis). 20
Supplement Figure 5: Sensitivity analysis subcortical volume effect sizes. 21
Supplement Figure 6: Cohen's d maps for Subcortical Shape analysis (Ventral view) 23
Supplement Figure 7: Covariance and overlap between surface and thickness at the vertex level 24
Supplement Figure 8: Comparison of effect sizes of CNVs on MRI-metric and disease-risk / cognition / #genes / pLI-sum. 26
Supplement Figure 9: Concordance Correlation Coefficient between subcortical volume, thickness, and surface effect sizes. 27
Supplement Figure 10: Correlation between Odds ratios for ASD/SZ, and Cohen's profiles' similarity of ASD/SZ with respective CNVs. 28
Supplement Figure 11: Latent dimensions across subcortical volumes of CNVs and NPDs. 29
Supplement Figure 12: Principal components analysis across vertex-wise Cohen's d maps of CNVs and NPDs (Ventral view). 30
Supplement Figure 13: Correlation between latent dimension of subcortical structures identified across volume, thickness, and surface. 32
Supplement Figure 14: CNV only principal components analysis across the subcortical volume, thickness, and surface. 33
Supplement Figure 15: Statistical significance of variance explained by PC1 + PC2. 34
Supplement Figure 16: Variance explained in Shape analysis versus number-of-vertices. 35
**References** 36

### MRI data acquisition and quality check

#### Participants

Clinically ascertained CNV carriers were recruited as either probands referred for genetic testing or as relatives. Controls were either non-carriers within the same families or individuals from the general population. We pooled data from 5 different cohorts: Cardiff University (Cardiff, UK), 16p11.2 European Consortium (Lausanne, Switzerland), University of Montreal (Canada), UCLA (Los Angeles, USA), and the Variation in Individuals Project (SVIP, USA). A subset of the participants with 16p11.2 proximal and 22q11.2 CNVs were included in prior publications (1–4). CNVs from non-clinical populations were identified in the UK Biobank (5, 6). PennCNV and QuantiSNP were used, with standard quality control metrics, to identify CNVs (7–9).

#### 16p11.2 European Consortium

MRI data of the EU participants were acquired on two 3T whole-body scanners. 14 carriers of a 16p11.2 proximal deletion and 17 duplication carriers, together with 59 controls (21 familial and 38 unrelated controls) were examined on a Magnetom TIM Trio (Siemens Healthcare, Erlangen, Germany), using a 12-channel RF receive head coil and RF body transmit coil. The remaining 16p11.2 proximal (13 deletions, 6 duplications), 1q21.2 distal (9 deletions, 7 duplications) carriers and controls (n=38) in the European cohort, were scanned on a Magnetom Prisma Syngo (Siemens Healthcare, Erlangen, Germany) using a 64-channel RF receive head coil and RF body transmit coil. T1-weighted (T1w) anatomical images acquired with the TIM Trio scanner used a Multi-Echo Magnetization Prepared RApid Gradient Echo sequence (ME-MPRAGE: 176 slices; 256×256 matrix; echo time (TE): TE1 = 1.64 ms, TE2 = 3.5 ms, TE3 = 5.36 ms, TE4 = 7.22 ms; repetition time (TR): 2530 ms; flip angle 7°). On the Prisma Syngo scanner, T1w images were acquired using a single-echo MPRAGE sequence (176 slices; 256×256 matrix; TE = 2.39 ms; TR = 2000 ms; flip angle 9°).

#### Simons Searchlight Consortium

Data were acquired using multi and single-echo sequences. 176 participants (38 del/ 34 dup 16p11.2 proximal carriers, 2 dup 1q21.1 distal carriers, and 102 familial controls) underwent the research MRI protocol at two imaging core sites on matched 3T Magnetom TIM Trio MRI scanners (Siemens Healthcare, Erlangen, Germany), using the vendor-supplied 32-channel phased-array radio-frequency head coils. 68 participants were scanned at University of California sites (UC) and 108 at the Children's Hospital of Philadelphia (CHOP). Structural MRI data included multi-echo T1w ME-MPRAGE using the following parameters: 176 slices, 256×256 matrix, TR = 2530 ms, TI = 1200 ms, TE = 1.64 ms, and flip angle 7°. Clinical MRI images (single-echo) obtained at the phenotyping core sites were also analyzed. The remaining 79 subjects (19 del/ 13 dup 16p11.2 proximal carriers, 12 del/8 dup 1q21.1 distal carriers and 27 familial controls) were scanned at the University of Washington Medical Center, Baylor University Medical Center, and Boston Children’s Hospital on two matched 3T Philips Achieva (Philips Healthcare, United States of America) and one unmatched Magnetom TIM Trio scanner (Siemens Healthcare, Erlangen, Germany), respectively. T1w images were acquired using a single-echo MPRAGE sequence and the following parameters: 160 slices; 256×256 matrix; TE = 2.98 ms; TR = 2300 ms; flip angle 9°. All multi-echo images were averaged following a Root-Mean Square (RMS) averaging method.

#### Brain Canada (BC, University of Montreal)

MRI scans for the Brain Canada cohort have been performed at the Montreal Neurological Institute with the same 3T scanner: Magnetom Prisma Syngo (Siemens Healthcare, Erlangen, Germany). Data included 16p11.2 proximal (3 deletions, 3 duplications), 1q21.2 distal (5 deletions, 1 duplication), 22q11.2 (1 duplication) carriers, and controls (n=26) T1w images were acquired using MPRAGE sequences, scanning protocol description is detailed on this website: http://www.bic.mni.mcgill.ca/users/jlewis/BrainCanada/MCIN/.

#### UCLA

Imaging data of 22q11.2 CNV carriers and typically developing (TD) controls were acquired at the University of California, Los Angeles (UCLA). Patients were ascertained from UCLA or Children's Hospital, Los Angeles Pediatric Genetics, Allergy/Immunology, and/or Craniofacial Clinics. We excluded 11 individuals from the analysis due to the insufficient quality of the imaging data (cf. Supplementary Methods, quality control). The final 22q11.2 sample includes 144 individuals (71 deletions, 19 duplications, and 54 controls). Demographically comparable TD comparison subjects were recruited from the same communities as patients via web-based advertisements and by posting flyers and brochures at local schools, pediatric clinics, and other community sites. Exclusion criteria for all study participants included significant neurological or medical conditions (unrelated to 22q11.2 mutation) that might affect brain structure, history of head injury with loss of consciousness, insufficient fluency in English, and/or substance or alcohol abuse or dependence within the past 6 months. The UCLA Institutional Review Board approved all study procedures and informed consent documents. Scanning was conducted on an identical 3 Tesla Siemens Trio MRI scanner with a 12-channel head coil at the University of California at Los Angeles Brain Mapping Center or at the Center for Cognitive Neuroscience.

#### Cardiff

Imaging acquisition in Cardiff was performed on a 3 T General Electric HDx MRI system (GE Medical Systems, Milwaukee, WI) using an eight-channel receive-only head RF coil. T1-weighted structural images were acquired with a 3D fast spoiled gradient echo (FSPGR) sequence (TR = 7.8 ms, TE = 3.0 ms, voxel size = 1 $mm^3$ isomorphic). Data included 1 16p11.2 proximal deletion, 1q21.2 distal (3 deletions, 1 duplication), 22q11.2 (3 deletions, 2 duplications) carriers and 15 controls.

#### MRI quality control

All MRI T1w NIfTI images were visually inspected by the same rater (CM) for head coverage, ghosting, and susceptibility artifacts. Images were also screened after segmentation to ensure good tissue classification accuracy. From the clinically ascertained dataset, 55 subjects were excluded for insufficient image quality or artifacts while from the non-clinically ascertained dataset 52 subjects were excluded following the same criteria. Quality assurance protocol for Freesurfer based cortical reconstructions led to the exclusion of an additional 34 scans. Numbers reported in Table 1 are after exclusion.

#### List of abbreviations

CNV: copy number variants; DEL: deletion; DUP: duplication; NPD: neurodevelopmental and psychiatric disorders; Corr: Pearson correlation; ASD: autism spectrum disorder; ADHD: attention deficit hyperactivity disorder; BD: bipolar disorder; MDD: major depressive disorder; OCD: obsessive-compulsive disorder; SZ: schizophrenia; PC: principal component; L: left hemisphere; Dim: dimension; ICV: Intracranial Volume; IQ: Intelligence Quotient; PCA: Principal components analysis.

### Supplementary Methods

##### *Subcortical volume and shape segmentation*

Raw data for all CNV carriers and controls were centrally preprocessed by the same investigators (CM, CC, and KK) using the same pipeline. Intracranial volume (ICV) was used as a global metric for total brain volume in all our analyses. For the subcortical volume analysis, we averaged the left and right hemisphere volumes for the 7 structures. For subcortical shape measures, we analyzed vertices across all 14 subcortical structures (bilateral), resulting in 27,000 vertices. Thickness is a proxy for subregional volume changes while the relationship between surface and volume depends on the local curvature of the region (31).

*Subcortical volume and shape Cohen's d maps from ENIGMA:*

Subcortical volume Cohen's *d* maps for the six neurodevelopmental and psychiatric disorders were obtained from previously published ENIGMA studies: schizophrenia (SZ (10)), major-depressive-disorder (MDD (11)), bipolar disorder (BD (12)), obsessive-compulsive-disorder (OCD (13)), autism-spectrum-disorder (ASD (14)), and attention-deficit-hyperactivity-disorder (ADHD (15)). The subcortical volume effect size maps are publicly available as part of the ENIGMA Toolbox (16). While volumetric Cohen's *d* maps were available for all 6 disorders in the ENIGMA Toolbox (10–15), Cohen's *d* maps for shape measures were only available for SZ and MDD (17, 18).

*Rationale for computing maximum effect sizes for subcortical shape metrics*

To estimate maximum effect sizes for shape metrics we averaged the absolute Cohen's *d* of the top decile across subcortical shape vertices. The rationale is that we assume that CNVs alter an unknown proportion of vertices with varying effect sizes. To provide a robust estimate, we chose to average vertices with Cohen's *d* in the top decile rather than taking the max Cohen's *d* which would result in a much noisier estimate. Because our power to detect vertices with significant differences (and properly estimate Cohen's *d*) is limited to those with the highest effect size we applied shrinkage across 90% of Cohen's *d*.

*Normative modeling*

We fitted Gaussian Processes Regression (GPR) model on controls and used age, sex, site, and ICV as covariates. Similar to (19, 20) we observed approximately linear effects of age for the accumbens, caudate, pallidum, and putamen, where the volumes peak during the first decade and decline over the years. Non-linear effects for age, flattened-inverted U-shape, were observed for ICV, amygdala, hippocampus, and thalamus (19).

##### *Site impact on Normative modeling*

To assess the impact of the site on Normative modeling, we used the leave-one-site-out approach, where Gaussian Processes modeling was run leaving data from either Brain-Canada or Cardiff sites (the 2 smallest samples) out, and estimating the effect sizes.

##### *Statistical testing of Cohen's d profiles (BrainSMASH)*

BrainSMASH (Brain Surrogate Maps with Autocorrelated Spatial Heterogeneity) was used for statistical testing of spatially autocorrelated Cohen's *d* profiles (brain maps). BrainSMASH simulates surrogate brain maps with spatial autocorrelation that is matched to spatial autocorrelation in a target brain map (21, 22). The spatial autocorrelation (SA) in brain maps is operationalized through the construction of a variogram, that quantifies, as a function of distance d, the variance between all pairs of points spatially separated by d. Further details are provided on the python package documentation page at: https://brainsmash.readthedocs.io/en/latest/approach.html.

##### *Covariance between shape metrics: Inter-individual maps of variation*

To estimate the maps of inter-individual variation between the local surface area and thickness we correlated the metrics at each vertex across all the controls. The metrics were adjusted for age, sex, site, and ICV, and Pearson's Correlation was computed between controls only. The correlations are then projected as vertex-wise measures to generate maps of covariance between local surface area and thickness and serve as maps of inter-individual variation between two metrics.

##### *Significance of the variance explained in Principal Component Analysis*

To test the significance of the variance explained in Principal Component Analysis, we generated null distribution of the variance explained using label shuffle (shuffling CNV and control labels) and re-ran PCA estimating the variance explained. The variance explained by PC1 + PC2 was then compared against the null distribution. For volume null distribution was generated using 1000 label shuffles. For Surface + Thickness, Surface, and Thickness null distribution was generated using 100 label shuffles.

##### *Impact of the number of vertices on variance explained*

To estimate the impact of the number of vertices or points, the granularity, on the variance explained in the Principal Component Analysis, we randomly sampled vertices and re-ran PCA estimating the variance explained. The variance explained was then plotted against an increasing number of vertices (sampled randomly). This allows us to observe the change in variance explained in PCA of shape metrics in the context of the level of granularity.

### Supplementary Tables

| CLINICAL ASCERTAINMENT | | | | | | | | | | |
|---|---|---|---|---|---|---|---|---|---|---|
| CNV loci | Copy number | Cohort | Age mean(SD) | Age mean(SD) | Male/Female | Male/Female | TIV mean(SD) | TIV mean(SD) | FSIQ mean(SD) | FSIQ mean(SD) |
| 1q21.1 | Deletions N=28 | EU N=9 | 29.43 (18.40) | 18.91 (12.91) | 15/13 | 8/1 | 1.22 (0.14) | 1.15 (0.09) | 90.85 (21.75) N=25 | 81.67 (21.57) N=9 |
| | | VIP N=11 | | 35.11 (20.66) | | 3/8 | | 1.27 (0.15) | | 99.33 (23.96) N=11 |
| | | BC N=5 | | 28.25 (18.46) | | 4/1 | | 1.17 (0.17) | | 87 (4.76) N=5 |
| | | Cardiff N=3 | | 40.18 (13.15) | | 0/3 | | 1.36 (0.02) | | (-) |
| | Duplications N=17 | EU N=6 | 34.29 (17.19) | 37.32 (19.58) | 9/8 | 4/2 | 1.57 (0.11) | 1.58 (0.07) | 95.56 (23.19) N=16 | 96.57 (11.59) N=6 |
| | | VIP N=9 | | 34.25 (15.84) | | 5/4 | | 1.54 (0.13) | | 93.80 (30.17) N=9 |
| | | BC N=1 | | 8.34 (-) | | 0/1 | | 1.57 (-) | | 106 (-) N=1 |
| | | Cardiff N=1 | | 39.45 (-) | | 0/1 | | 1.76 (-) | | (-) |
| 16p11.2 | Deletions N=78 | EU N=24 | 17.14 (11.97) | 21.23 (13.51) | 34/44 | 12/12 | 1.54 (0.17) | 1.44 (0.14) | 82.17 (14.99) N=64 | 74.38 (14.61) N=13 |
| | | VIP N=50 | | 13.82 (9.47) | | 20/30 | | 1.58 (0.17) | | 83.98 (14.36) N=48 |
| | | BC N=3 | | 31.64 (10.26) | | 1/2 | | 1.46 (0.09) | | 87 (21) N=3 |
| | | Cardiff N=1 | | 43.04 (-) | | 1/0 | | 2.01 (-) | | (-) |
| | Duplications N=68 | EU N=21 | 31.01(14.91) | 32.55 (13.20) | 38/30 | 11/10 | 1.33 (0.17) | 1.36 (0.16) | 85 (19.70) N=63 | 72.71 (16.30) N=17 |
| | | VIP N=44 | | 30.52 (15.30) | | 24/20 | | 1.30 (0.15) | | 90.44 (18.49) N=43 |
| | | BC N=3 | | 26.89 (25.29) | | 3/0 | | 1.49 (0.42) | | 86.67 (23.50) N=3 |
| 22q11.2 | Deletions N=68 | UCLA N=65 | 16.35 (8.56) | 15.66 (7.24) | 33/35 | 32/33 | 1.30 (0.15) | 1.30 (0.15) | 77.41 (13.51) N=48 | 77.41 (13.51) N=48 |
| | | Cardiff N=3 | | 32.56 (20.60) | | 1/2 | | 1.37 (0.11) | | (-) |
| | Duplications N=19 | UCLA N=16 | 19.66 (14.24) | 17.32 (12.51) | 8/11 | 7/9 | 1.47 (0.16) | 1.45 (0.17) | 97.83 (20.34) N=12 | 99.18 (20.76) N=11 |
| | | BC N=1 | | 13.67 (-) | | 0/1 | | 1.50 (-) | | 83 (-) N=1 |
| | | Cardiff N=2 | | 44.94 (4.77) | | 1/1 | | 1.55 (0.16) | | (-) |
| Controls N=317 | | EU N=96 | 25.91 (14.57) | 30.33 (12.95) | 182/135 | 62/34 | 1.46 (0.15) | 1.49 (0.14) | 106.73 (15.03) N=224 | 102.15 (12.98) N=59 |
| | | VIP N=127 | | 24.03 (14.56) | | 70/57 | | 1.44 (0.14) | | 108.88 (11.82) N=90 |
| | | UCLA N=46 | | 13.37 (4.96) | | 26/20 | | 1.39 (0.13) | | 112.43 (20.95) N=44 |
| | | BC N=33 | | 33.56 (14.90) | | 17/16 | | 1.52 (0.17) | | 101.11 (13.18) N=31 |
| | | Cardiff N=15 | | 40.18 (11.12) | | 7/8 | | 1.54 (0.15) | | (-) |

| NON-CLINICAL ASCERTAINMENT | | | | | | |
|---|---|---|---|---|---|---|
| CNV loci | Copy number | Cohort | Age mean(SD) | Male/Female | TIV mean(SD) | UKB FI mean(SD) |
| 1q21.1 | Deletions N=12 | UKBB | 59.11 (6.71) | 7/5 | 1.35 (0.12) | -0.8 (0.5) N=10 |
| | Duplications N=13 | | 60.56 (7) | 4/9 | 1.55 (0.14) | 0.2 (1.3) N=11 |
| TAR | Duplications N=31 | | 60 (8) | 14/17 | 1.48 (0.16) | -0.1 (1.2) N=27 |
| 13q12.12 | Duplications N=21 | | 62 (8) | 11/10 | 1.54 (0.15) | 0.1 (1.2) N=18 |
| 15q11.2 | Deletions N=108 | | 65 (7.10) | 59/49 | 1.54 (0.15) | -0.3 (0.9) N=99 |
| | Duplications N=144 | | 64 (7.31) | 77/67 | 1.49 (0.15) | 0 (1.1) N=132 |
| 16p11.2 | Deletions N=4 | | 65.6 (3.2) | 3/1 | 1.56 (0.13) | 0.8 (0.5) N=2 |
| | Duplications N=7 | | 69.3 (2.1) | 2/5 | 1.29 (0.11) | -1.6 (0.2) N=5 |
| 16p13.11 | Duplications N=50 | | 66 (6) | 26/24 | 1.49 (0.17) | -0.2 (1.2) N=46 |
| 22q11.2 | Duplications N=7 | | 62 (9.5) | 3/4 | 1.55 (0.17) | -0.2 (1.1) N=6 |
| Controls N=465 | | | 62.13 (7.40) | 205/260 | 1.51 (0.16) | 0 (1) N=445 |

#### Supplement Table 1: Detailed demographics

Legend: Detailed demographics and cohort information. EU: 16p11.2 European Consortium, VIP: Simons Searchlight Consortium, BC: Brain Canada, CNV: Copy Number Variant, SD: Standard deviation, TIV: total intracranial volume, FSIQ: Full-scale IQ, UKB FI: UK Biobank fluid intelligence. CNV carriers and controls from the clinically ascertained group come from 5 different cohorts, while non-clinically ascertained participants were identified in the UK Biobank. UK Biobank fluid intelligence scores (UKB field:20016) were adjusted for age, age2, sex, site, and then z-scored. Part of the data was previously published in (2, 23) and (1, 24).

| Loci | Type | nGenes | Effect sizes | | | | | |
|---|---|---|---|---|---|---|---|---|
| | | | IQ_loss | OR ASD/SZ | ICV | Volume | Thickness | Surface |
| 1q21.1 | Del | 7 | 15 | 6.4 | 2.09 | 0.36 | 0.68 | 0.79 |
| | Dup | 7 | 25 | 5.3 | 0.72 | 0.81 | 0.62 | 0.6 |
| 16p11.2 | Del | 27 | 26 | 14.3 | 0.52 | 0.84 | 0.89 | 1.07 |
| | Dup | 27 | 11 | 11.7 | 1.25 | 0.64 | 0.68 | 1.02 |
| 22q11.2 | Del | 49 | 28.8 | 32.3 | 0.43 | 0.92 | 1.03 | 1.24 |
| | Dup | 42 | 8.3 | 2 | 0.24 | 0.63 | 0.85 | 0.72 |
| 15q11.2 | Del | 4 | 3 | 1.3 | 0.18 | 0.37 | 0.44 | 0.53 |
| | Dup | 4 | 0.9 | 1.8 | 0.31 | 0.21 | 0.36 | 0.37 |
| 16p13.11 | Dup | 6 | 2 | 1.5 | 0.14 | 0.36 | 0.66 | 0.54 |
| 13q12.12 | Dup | 5 | 0.6 | - | 0.23 | 0.74 | 0.68 | 0.65 |
| TAR | Dup | 15 | 2.4 | 1 | 0.19 | 0.62 | 0.65 | 0.52 |
| ASD | ENIGMA NPDs | - | - | - | 0.13 | 0.13 | - | - |
| ADHD | | - | - | - | 0.1 | 0.19 | - | - |
| BD | | - | - | - | 0 | 0.23 | - | - |
| MDD | | - | - | - | 0.03 | 0.14 | 0.12 | 0.11 |
| OCD | | - | - | - | 0.01 | 0.16 | - | - |
| SZ | | - | - | - | 0.12 | 0.46 | 0.39 | 0.34 |

#### Supplement Table 2: Summary of effect sizes across CNVs and NPDs, for MRI-metrics.

Legend: Summary effect sizes for IQ-loss/disease-risk, subcortical volumes, thickness, surface, and ICV. Thickness represents local radial distance, and surface (Jacobian) represents local surface area dilation/contraction. OR: Odds ratio; ICV: Intracranial volume;

Del: deletion; Dup: Duplication; ASD: autism spectrum disorder; ADHD: attention deficit hyperactivity disorder; BD: bipolar disorder; MDD: major depressive disorder; OCD: obsessive-compulsive disorder; SZ: schizophrenia; IQ: Intelligence Quotient.

| Comparison | | Welch Two Sample t-test | | | | | Pearson | | Concordance Correlation Coefficient (CCC) | | | |
|---|---|---|---|---|---|---|---|---|---|---|---|---|
| ES1 | ES2 | MeanES1 | MeanES2 | DF | t-stat | Pval | Corr | CorrPval | CCC_rho | 95% CI [low high] | | Bias Factor |
| ICV | SubCortVol | 0.573 | 0.591 | 12.972 | -0.094 | 0.927 | -0.095 | 0.78 | -0.064 | -0.466 | 0.359 | 0.676 |
| ICV | Thick | 0.573 | 0.685 | 11.986 | -0.595 | 0.563 | 0.063 | 0.853 | 0.035 | -0.316 | 0.378 | 0.556 |
| ICV | Surf | 0.573 | 0.732 | 13.937 | -0.802 | 0.436 | 0.335 | 0.315 | 0.236 | -0.209 | 0.6 | 0.706 |
| SubCortVol | Thick | 0.591 | 0.685 | 19.193 | -1.042 | 0.31 | 0.766 | **0.006** | 0.676 | 0.247 | 0.884 | 0.883 |
| SubCortVol | Surf | 0.591 | 0.732 | 19.567 | -1.305 | 0.207 | 0.68 | **0.021** | 0.575 | 0.098 | 0.837 | 0.845 |
| Thick | Surf | 0.732 | 0.685 | 17.891 | 0.464 | 0.648 | 0.833 | **0.001** | 0.766 | 0.437 | 0.914 | 0.919 |

#### Supplement Table 3: Concordance between CNV effect size across metrics: ICV-volume-surface-thickness.

Legend: Welch t-test comparing the effect size across SubCortical measures and ICV reported in Supplement Table 1. ICV: Intracranial volume; SubCortVol: subcortical volume; Thick: thickness shape metric; Surf: local surface area shape metric; ES: effect size; MeanES1: mean effect size for metric 1; MeanES2: mean effect size for metric2; DF: degree of freedom; t-stat: T-statistic; Pval: p-value; Corr: correlation; CorrPval: Correlation p-value; CCC: Concordance correlation coefficient; CI: confidence interval;

| ROIs | 1q21.1 | | 16p11.2 | | 22q11.2 | | 15q11.2 | | 16p13.11 | 13q12.12 | TAR | Average | Total Vertices |
|---|---|---|---|---|---|---|---|---|---|---|---|---|---|
| | Del | Dup | Del | Dup | Del | Dup | Del | Dup | Dup | Dup | Dup | | |
| Lthal | 521 | 231 | 1074 | 578 | 1006 | 190 | 41 | 50 | 42 | 25 | 17 | 343.1818 | 2502 |
| Lcaud | 33 | 246 | 906 | 934 | 1094 | 346 | 286 | 498 | 525 | 92 | 176 | 466.9091 | 2502 |
| Lput | 83 | 59 | 1203 | 966 | 1136 | 163 | 571 | 530 | 65 | 3 | 16 | 435.9091 | 2502 |
| Lpal | 214 | 30 | 455 | 209 | 137 | 13 | 95 | 264 | 85 | 26 | 3 | 139.1818 | 1254 |
| Lhippo | 326 | 234 | 298 | 305 | 1193 | 16 | 79 | 67 | 122 | 46 | 60 | 249.6364 | 2502 |
| Lamyg | 135 | 0 | 316 | 284 | 483 | 228 | 173 | 302 | 88 | 21 | 25 | 186.8182 | 1368 |
| Laccumb | 97 | 1 | 303 | 181 | 662 | 131 | 10 | 169 | 28 | 5 | 263 | 168.1818 | 930 |
| Rthal | 489 | 175 | 838 | 578 | 815 | 132 | 51 | 63 | 58 | 0 | 50 | 295.3636 | 2502 |
| Rcaud | 102 | 74 | 629 | 643 | 1433 | 143 | 128 | 634 | 420 | 236 | 7 | 404.4545 | 2502 |
| Rput | 243 | 70 | 823 | 735 | 1112 | 53 | 456 | 261 | 68 | 47 | 54 | 356.5455 | 2502 |
| Rpal | 199 | 1 | 458 | 377 | 257 | 23 | 345 | 485 | 11 | 0 | 0 | 196 | 1254 |
| Rhippo | 608 | 71 | 504 | 669 | 1545 | 215 | 112 | 177 | 196 | 36 | 142 | 388.6364 | 2502 |
| Ramyg | 168 | 130 | 756 | 602 | 742 | 393 | 139 | 164 | 351 | 71 | 86 | 327.4545 | 1368 |
| Raccumb | 37 | 98 | 101 | 148 | 595 | 66 | 93 | 8 | 0 | 0 | 1 | 104.2727 | 930 |
| Average | 232.5 | 101.43 | 618.86 | 514.93 | 872.14 | 150.86 | 184.21 | 262.29 | 147.071 | 43.42857 | 64.2857 | | 27120 |

#### Supplement Table 4: Number of significant vertices per ROI across 11 CNVs for thickness.

Legend: Number of Vertices surviving FDR correction (<0.05) applied across all vertices of 11 CNVs for subcortical shape analysis of thickness. The largest average value across CNVs and across ROIs is highlighted in yellow. Cells in red show a higher count. Corresponding maps are shown in **Figure 2B**. Abbreviations, DEL: deletion; DUP: duplication; ACC: accumbens; AMY: amygdala; CAUD: caudate; HIP: hippocampus; PUT: putamen; PAL: pallidum; THAL: thalamus; L: left hemisphere; R: right hemisphere.

| ROIs | 1q21.1 | | 16p11.2 | | 22q11.2 | | 15q11.2 | | 16p13.11 | 13q12.12 | TAR | Average | Total Vertices |
|---|---|---|---|---|---|---|---|---|---|---|---|---|---|
| | Del | Dup | Del | Dup | Del | Dup | Del | Dup | Dup | Dup | Dup | | |
| Lthal | 0.550 | 0.622 | 0.574 | 0.445 | 0.533 | 0.626 | 0.307 | 0.287 | 0.452 | 0.631 | 0.517 | 0.504 | 2502 |
| Lcaud | 0.483 | 0.583 | 0.433 | 0.474 | 0.587 | 0.667 | 0.328 | 0.300 | 0.559 | 0.640 | 0.540 | 0.509 | 2502 |
| Lput | 0.495 | 0.552 | 0.538 | 0.437 | 0.692 | 0.632 | 0.370 | 0.303 | 0.434 | 0.621 | 0.538 | 0.510 | 2502 |
| Lpal | 0.606 | 0.574 | 0.454 | 0.445 | 0.429 | 0.618 | 0.317 | 0.308 | 0.448 | 0.685 | 0.550 | 0.494 | 1254 |
| Lhippo | 0.590 | 0.574 | 0.464 | 0.408 | 0.619 | 0.702 | 0.312 | 0.266 | 0.438 | 0.671 | 0.590 | 0.512 | 2502 |
| Lamyg | 0.523 | 0.000 | 0.447 | 0.396 | 0.519 | 0.652 | 0.345 | 0.306 | 0.457 | 0.635 | 0.513 | 0.436 | 1368 |
| Laccumb | 0.529 | 0.528 | 0.417 | 0.382 | 0.529 | 0.665 | 0.285 | 0.282 | 0.421 | 0.626 | 0.586 | 0.477 | 930 |
| Rthal | 0.516 | 0.612 | 0.560 | 0.459 | 0.560 | 0.645 | 0.313 | 0.266 | 0.440 | 0.000 | 0.538 | 0.446 | 2502 |
| Rcaud | 0.504 | 0.548 | 0.453 | 0.476 | 0.643 | 0.648 | 0.327 | 0.297 | 0.566 | 0.683 | 0.529 | 0.516 | 2502 |
| Rput | 0.534 | 0.549 | 0.572 | 0.456 | 0.604 | 0.602 | 0.361 | 0.284 | 0.461 | 0.640 | 0.553 | 0.511 | 2502 |
| Rpal | 0.542 | 0.535 | 0.440 | 0.515 | 0.474 | 0.614 | 0.352 | 0.304 | 0.450 | 0.000 | 0.000 | 0.384 | 1254 |
| Rhippo | 0.576 | 0.578 | 0.419 | 0.458 | 0.669 | 0.644 | 0.334 | 0.285 | 0.442 | 0.747 | 0.665 | 0.529 | 2502 |
| Ramyg | 0.512 | 0.621 | 0.592 | 0.457 | 0.635 | 0.694 | 0.325 | 0.278 | 0.489 | 0.664 | 0.557 | 0.530 | 1368 |
| Raccumb | 0.498 | 0.587 | 0.397 | 0.374 | 0.542 | 0.601 | 0.320 | 0.269 | 0.000 | 0.592 | 0.500 | 0.425 | 930 |
| Average | 0.533 | 0.533 | 0.483 | 0.442 | 0.574 | 0.643 | 0.328 | 0.288 | 0.433 | 0.560 | 0.513 | | 27120 |

#### Supplement Table 5: Mean of absolute Cohen's *d* for significant vertices per ROI across 11 CNVs for thickness.

Legend: Mean of absolute Cohen's *d* for vertices surviving FDR correction (<0.05) applied across all vertices of 11 CNVs for subcortical shape analysis of thickness. Corresponding maps are shown in **Figure 2B**. Abbreviations, DEL: deletion; DUP: duplication; ACC: accumbens; AMY: amygdala; CAUD: caudate; HIP: hippocampus; PUT: putamen; PAL: pallidum; THAL: thalamus; L: left hemisphere; R: right hemisphere.

| ROIs | 1q21.1 | | 16p11.2 | | 22q11.2 | | 15q11.2 | | 16p13.11 | 13q12.12 | TAR | Average | Total Vertices |
|---|---|---|---|---|---|---|---|---|---|---|---|---|---|
| | Del | Dup | Del | Dup | Del | Dup | Del | Dup | Dup | Dup | Dup | | |
| Lthal | 591 | 93 | 1389 | 1703 | 1261 | 189 | 350 | 30 | 3 | 118 | 71 | 527.0909 | 2502 |
| Lcaud | 26 | 118 | 1495 | 1412 | 742 | 195 | 287 | 97 | 167 | 18 | 50 | 418.8182 | 2502 |
| Lput | 58 | 13 | 1101 | 1289 | 897 | 208 | 1009 | 709 | 49 | 10 | 0 | 485.7273 | 2502 |
| Lpal | 311 | 4 | 203 | 367 | 72 | 62 | 266 | 281 | 104 | 0 | 6 | 152.3636 | 1254 |
| Lhippo | 320 | 343 | 552 | 1117 | 1522 | 59 | 253 | 382 | 265 | 166 | 2 | 452.8182 | 2502 |
| Lamyg | 155 | 1 | 232 | 252 | 564 | 151 | 410 | 445 | 120 | 44 | 33 | 218.8182 | 1368 |
| Laccumb | 8 | 20 | 518 | 1 | 785 | 126 | 26 | 74 | 15 | 0 | 32 | 145.9091 | 930 |
| Rthal | 367 | 3 | 1262 | 1647 | 970 | 32 | 137 | 4 | 5 | 0 | 38 | 405.9091 | 2502 |
| Rcaud | 15 | 321 | 1184 | 670 | 1339 | 14 | 240 | 346 | 70 | 26 | 34 | 387.1818 | 2502 |
| Rput | 327 | 13 | 743 | 1153 | 792 | 29 | 1022 | 405 | 36 | 0 | 52 | 415.6364 | 2502 |
| Rpal | 263 | 2 | 455 | 584 | 181 | 1 | 496 | 438 | 11 | 0 | 8 | 221.7273 | 1254 |
| Rhippo | 591 | 225 | 1220 | 1600 | 1920 | 63 | 297 | 170 | 220 | 100 | 40 | 586 | 2502 |
| Ramyg | 76 | 55 | 755 | 607 | 812 | 364 | 293 | 185 | 418 | 157 | 99 | 347.3636 | 1368 |
| Raccumb | 0 | 24 | 339 | 48 | 850 | 62 | 144 | 30 | 4 | 3 | 8 | 137.4545 | 930 |
| Average | 222 | 88.21 | 817.7 | 889.3 | 907.6 | 111.1 | 373.6 | 256.9 | 106.214 | 45.85714 | 33.7857 | | 27120 |

Supplement Table 6: Number of significant vertices per ROI across 11 CNVs for local surface area.

Legend: Number of vertices surviving FDR correction (<0.05) applied across all vertices of 11 CNVs for subcortical shape analysis of the local surface area metric (Log-Jacobian). Cells in red show a higher count. The largest average value across CNVs and across ROIs is highlighted in yellow. Corresponding maps are shown in **Figure 2A**.

Abbreviations, DEL: deletion; DUP: duplication; ACC: accumbens; AMY: amygdala; CAUD: caudate; HIP: hippocampus; PUT: putamen; PAL: pallidum; THAL: thalamus; L: left hemisphere; R: right hemisphere.

| ROIs | 1q21.1 | | 16p11.2 | | 22q11.2 | | 15q11.2 | | 16p13.11 | 13q12.12 | TAR | Average | Total Vertices |
|---|---|---|---|---|---|---|---|---|---|---|---|---|---|
| | Del | Dup | Del | Dup | Del | Dup | Del | Dup | Dup | Dup | Dup | | |
| Lthal | 0.622 | 0.540 | 0.619 | 0.545 | 0.587 | 0.586 | 0.338 | 0.270 | 0.394 | 0.636 | 0.536 | 0.516 | 2502 |
| Lcaud | 0.489 | 0.583 | 0.472 | 0.522 | 0.575 | 0.579 | 0.323 | 0.290 | 0.441 | 0.622 | 0.530 | 0.493 | 2502 |
| Lput | 0.482 | 0.515 | 0.472 | 0.508 | 0.569 | 0.593 | 0.401 | 0.315 | 0.412 | 0.608 | 0.000 | 0.443 | 2502 |
| Lpal | 0.547 | 0.518 | 0.411 | 0.434 | 0.410 | 0.613 | 0.345 | 0.302 | 0.438 | 0.000 | 0.519 | 0.413 | 1254 |
| Lhippo | 0.493 | 0.570 | 0.466 | 0.487 | 0.681 | 0.601 | 0.327 | 0.274 | 0.470 | 0.638 | 1.043 | 0.550 | 2502 |
| Lamyg | 0.507 | 0.495 | 0.378 | 0.395 | 0.480 | 0.642 | 0.380 | 0.288 | 0.446 | 0.628 | 0.526 | 0.470 | 1368 |
| Laccumb | 0.443 | 0.516 | 0.510 | 0.324 | 0.479 | 0.613 | 0.297 | 0.267 | 0.421 | 0.000 | 0.504 | 0.398 | 930 |
| Rthal | 0.550 | 0.510 | 0.569 | 0.658 | 0.536 | 0.579 | 0.319 | 0.257 | 0.398 | 0.000 | 0.553 | 0.448 | 2502 |
| Rcaud | 0.455 | 0.586 | 0.481 | 0.516 | 0.592 | 0.584 | 0.360 | 0.291 | 0.430 | 0.619 | 0.519 | 0.494 | 2502 |
| Rput | 0.539 | 0.527 | 0.448 | 0.535 | 0.498 | 0.556 | 0.405 | 0.290 | 0.417 | 0.584 | 0.527 | 0.484 | 2502 |
| Rpal | 0.531 | 0.513 | 0.451 | 0.538 | 0.539 | 0.543 | 0.363 | 0.296 | 0.428 | 0.000 | 0.518 | 0.429 | 1254 |
| Rhippo | 0.543 | 0.573 | 0.503 | 0.614 | 0.851 | 0.597 | 0.318 | 0.284 | 0.460 | 0.635 | 0.565 | 0.540 | 2502 |
| Ramyg | 0.507 | 0.528 | 0.504 | 0.424 | 0.648 | 0.640 | 0.338 | 0.274 | 0.467 | 0.644 | 0.523 | 0.500 | 1368 |
| Raccumb | 0.000 | 0.552 | 0.491 | 0.369 | 0.612 | 0.585 | 0.318 | 0.267 | 0.388 | 0.598 | 0.506 | 0.426 | 930 |
| Average | 0.479 | 0.538 | 0.484 | 0.491 | 0.575 | 0.594 | 0.345 | 0.283 | 0.429 | 0.444 | 0.526 | | 27120 |

#### Supplement Table 7: Mean of absolute Cohen's *d* for significant vertices per ROI across 11 CNVs for local surface area.

Legend: Mean of absolute Cohen's *d* for vertices surviving FDR correction (<0.05) applied across all vertices of 11 CNVs for subcortical shape analysis of the local surface area metric (Log-Jacobian). Corresponding maps are shown in **Figure 2A**. Abbreviations, DEL: deletion; DUP: duplication; ACC: accumbens; AMY: amygdala; CAUD: caudate; HIP: hippocampus; PUT: putamen; PAL: pallidum; THAL: thalamus; L: left hemisphere; R: right hemisphere.

| Surf-Thick | ROIs | 1q21.1 | | 16p11.2 | | 22q11.2 | | 15q11.2 | | 16p13.11 | 13q12.12 | TAR |
|---|---|---|---|---|---|---|---|---|---|---|---|---|
| | | Del | Dup | Del | Dup | Del | Dup | Del | Dup | Dup | Dup | Dup |
| Overlap: Both Positive | Lthal | 0 | 173 | 1514 | 7 | 30 | 60 | 1 | 3 | 1 | 144 | 0 |
| | Lcaud | 1 | 122 | 800 | 31 | 682 | 139 | 12 | 32 | 489 | 131 | 229 |
| | Lput | 36 | 3 | 597 | 0 | 610 | 262 | 0 | 0 | 138 | 17 | 16 |
| | Lpal | 0 | 24 | 193 | 0 | 117 | 72 | 0 | 0 | 185 | 4 | 4 |
| | Lhippo | 142 | 0 | 367 | 33 | 8 | 66 | 0 | 0 | 255 | 190 | 9 |
| | Lamyg | 112 | 0 | 496 | 0 | 29 | 339 | 14 | 0 | 209 | 87 | 81 |
| | Laccumb | 39 | 14 | 0 | 0 | 842 | 265 | 5 | 0 | 21 | 0 | 302 |
| | Rthal | 110 | 153 | 1197 | 22 | 712 | 87 | 23 | 10 | 4 | 0 | 0 |
| | Rcaud | 9 | 272 | 517 | 84 | 1323 | 34 | 12 | 82 | 431 | 227 | 55 |
| | Rput | 89 | 14 | 339 | 0 | 576 | 41 | 0 | 0 | 105 | 52 | 117 |
| | Rpal | 0 | 8 | 265 | 0 | 183 | 5 | 4 | 0 | 15 | 0 | 11 |
| | Rhippo | 114 | 7 | 878 | 6 | 11 | 147 | 0 | 0 | 375 | 91 | 110 |
| | Ramyg | 1 | 169 | 851 | 0 | 17 | 641 | 0 | 0 | 657 | 249 | 206 |
| | Raccumb | 24 | 147 | 8 | 0 | 865 | 127 | 0 | 0 | 13 | 7 | 13 |
| Overlap: Both Negative | Lthal | 1003 | 15 | 49 | 1231 | 1184 | 208 | 229 | 90 | 38 | 0 | 119 |
| | Lcaud | 51 | 141 | 143 | 1491 | 289 | 340 | 411 | 461 | 67 | 1 | 0 |
| | Lput | 85 | 40 | 590 | 1484 | 177 | 23 | 1252 | 929 | 18 | 0 | 0 |
| | Lpal | 361 | 0 | 129 | 509 | 38 | 0 | 343 | 469 | 0 | 20 | 5 |
| | Lhippo | 209 | 562 | 53 | 920 | 1216 | 13 | 211 | 389 | 14 | 0 | 0 |
| | Lamyg | 91 | 4 | 0 | 498 | 590 | 0 | 436 | 665 | 0 | 0 | 0 |
| | Laccumb | 32 | 0 | 509 | 170 | 0 | 0 | 42 | 219 | 0 | 1 | 0 |
| | Rthal | 402 | 0 | 54 | 1077 | 254 | 60 | 127 | 44 | 58 | 0 | 86 |
| | Rcaud | 81 | 7 | 71 | 765 | 180 | 80 | 320 | 446 | 5 | 9 | 0 |
| | Rput | 298 | 65 | 434 | 1228 | 209 | 3 | 1170 | 566 | 0 | 0 | 0 |
| | Rpal | 413 | 2 | 86 | 779 | 42 | 5 | 702 | 788 | 0 | 0 | 0 |
| | Rhippo | 342 | 288 | 44 | 1164 | 1548 | 107 | 280 | 310 | 0 | 0 | 0 |
| | Ramyg | 176 | 15 | 75 | 922 | 875 | 0 | 376 | 358 | 0 | 0 | 0 |
| | Raccumb | 7 | 0 | 317 | 209 | 0 | 0 | 229 | 44 | 0 | 0 | 0 |

#### Supplement Table 8: Number of surface and thickness overlap vertices per ROI across 11 CNVs.

Legend: Overlap between surface and thickness vertices for each CNV; significant for either metric or for both. The number of vertices, where both metrics are positive (top half) or negative (bottom half), for 11 CNVs. Cells in red show a higher count. Corresponding maps are shown in **Supplement Figure 7**.

| ROIs | Surf-Thick PC1 Overlap | | Surf-Thick PC2 Overlap | |
|---|---|---|---|---|
| | Both Positive | Both Negative | Both Positive | Both Negative |
| Lthal | 126 | 1528 | 2423 | 8 |
| Lcaud | 920 | 479 | 424 | 784 |
| Lput | 1073 | 566 | 217 | 1139 |
| Lpal | 760 | 69 | 327 | 392 |
| Lhippo | 262 | 1059 | 508 | 782 |
| Lamyg | 248 | 550 | 55 | 955 |
| Laccumb | 642 | 9 | 0 | 699 |
| Rthal | 892 | 326 | 2250 | 13 |
| Rcaud | 1505 | 291 | 647 | 629 |
| Rput | 932 | 638 | 60 | 1458 |
| Rpal | 481 | 281 | 176 | 361 |
| Rhippo | 138 | 1250 | 668 | 761 |
| Ramyg | 32 | 887 | 52 | 1086 |
| Raccumb | 795 | 0 | 92 | 361 |

Supplement Table 9: Number of surface and thickness PC overlap vertices per ROI.

Legend: Overlap between PC1 and PC2 of surface and thickness vertices. The number of vertices, where both metrics are positive or negative, for PC1 and PC2 respectively. Cells in red show a higher count. Corresponding maps are shown in **Figure 4E**.

### Supplementary Figures

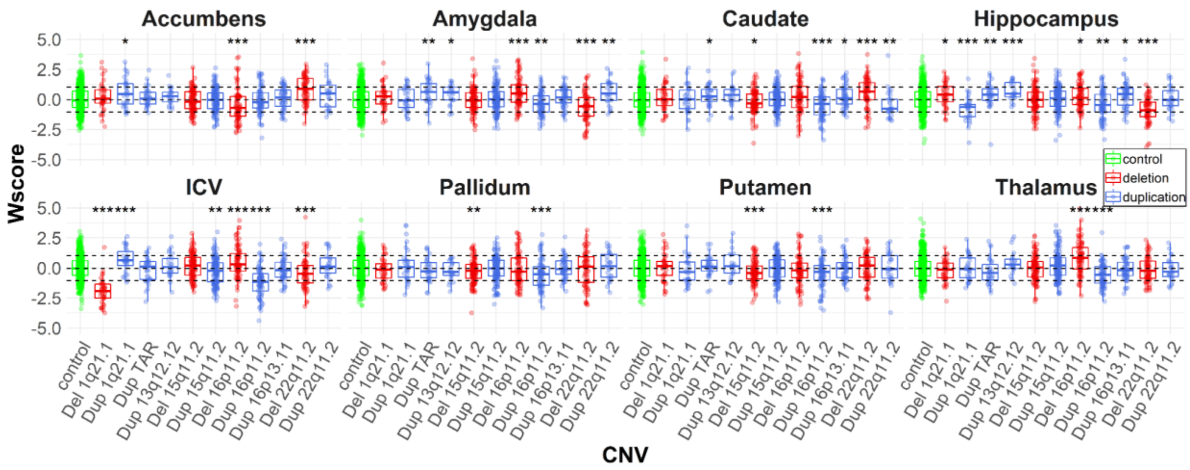


Supplement Figure 1: W-score obtained using Gaussian Processes modeling.

Legend: Boxplots showing the distribution of ICV and SubCortical W-scores (Z Score using mean and sigma of controls based on Gaussian Processes modeling) for CNVs. Gaussian Processes are modeled using age, sex, site, and ICV (except for ICV) as covariates. P-values for comparison of CNV W-scores w.r.t control W-scores are shown with symbols. Significant estimates with nominal p-value <0.05: “*”, p-value <0.01: “**”, and p-value <0.001: “***”.

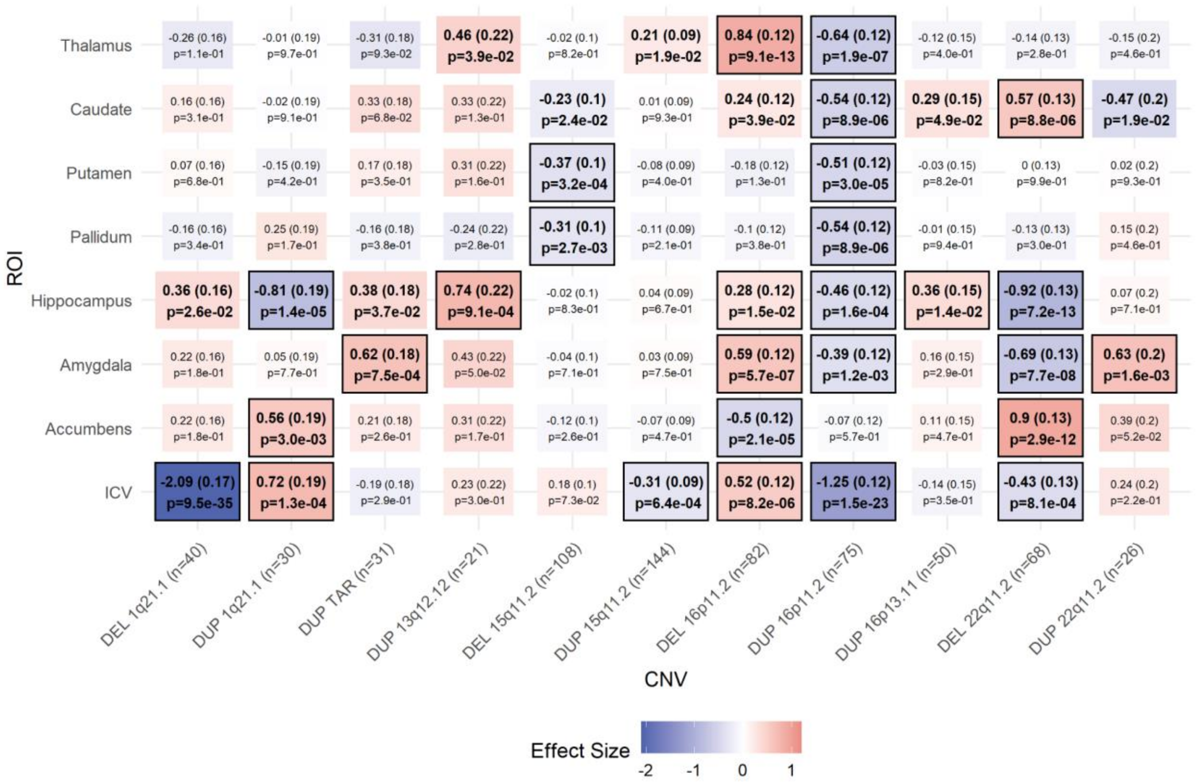

Supplement Figure 2: Effect sizes, SE, and p-values for subcortical volumes and ICV.

Legend: Tile plot showing the Cohen's *d* (SE) and p-values for SubCortical volumes and ICV. Case-control differences are calculated (lm in R) using W-scores from Gaussian processes regression (GPR, with age, sex, site, and ICV as covariates). Significant Cohen's *d* with nominal p-value <0.05 are in bold, and FDR p-value <0.05 are shown with rectangles. Darker color represents higher magnitudes. lh and rh denote the left and right hemispheres respectively. DEL: deletions; DUP: duplications.

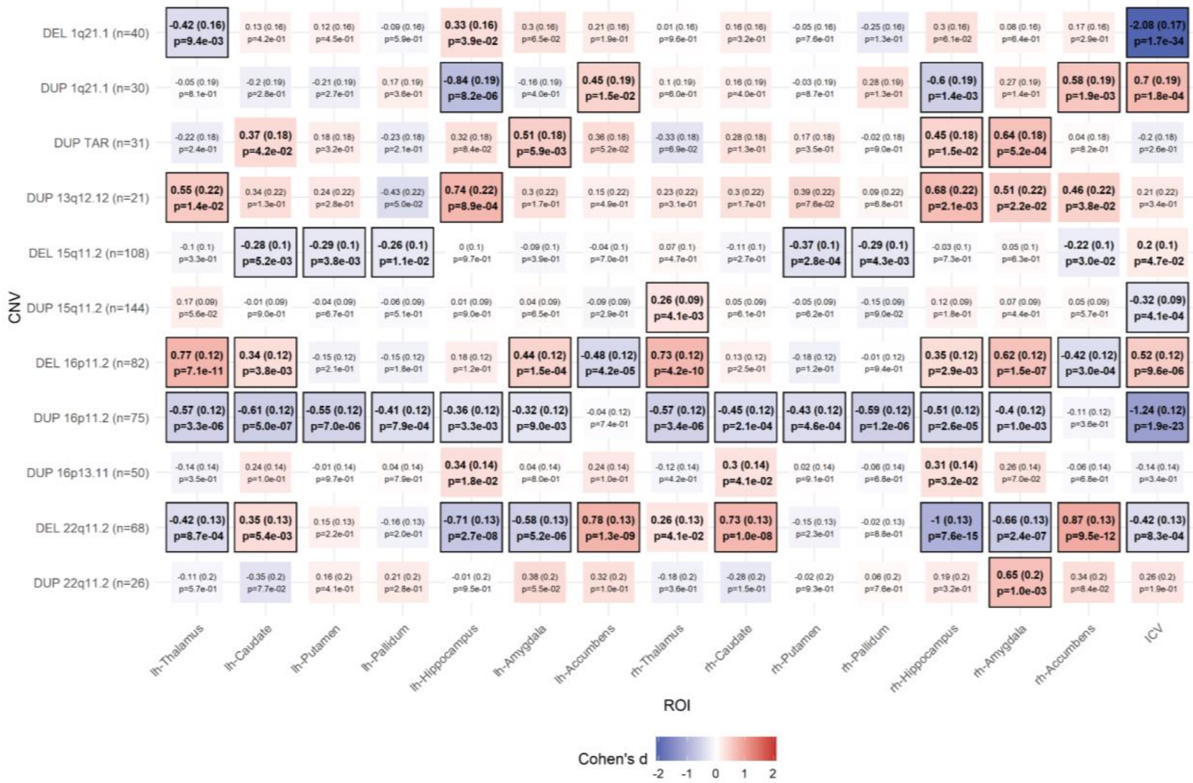


Supplement Figure 3: Sensitivity analysis- bilateral effect sizes (Cohen's *d*) for subcortical structures and ICV.

Cohen's *d* (SE) and p-values for subcortical structures for CNVs. Case-control differences are calculated (lm in R) using W-scores obtained from Gaussian processes regression (GPR, with age, sex, site, and ICV as covariates). Significant Cohen**'s** *d* with nominal p-value <0.05 are in bold, and FDR p-value <0.05 are shown with rectangles. Darker color represents higher magnitudes. lh and rh denote the left and right hemispheres respectively. DEL: deletions; DUP: duplications.

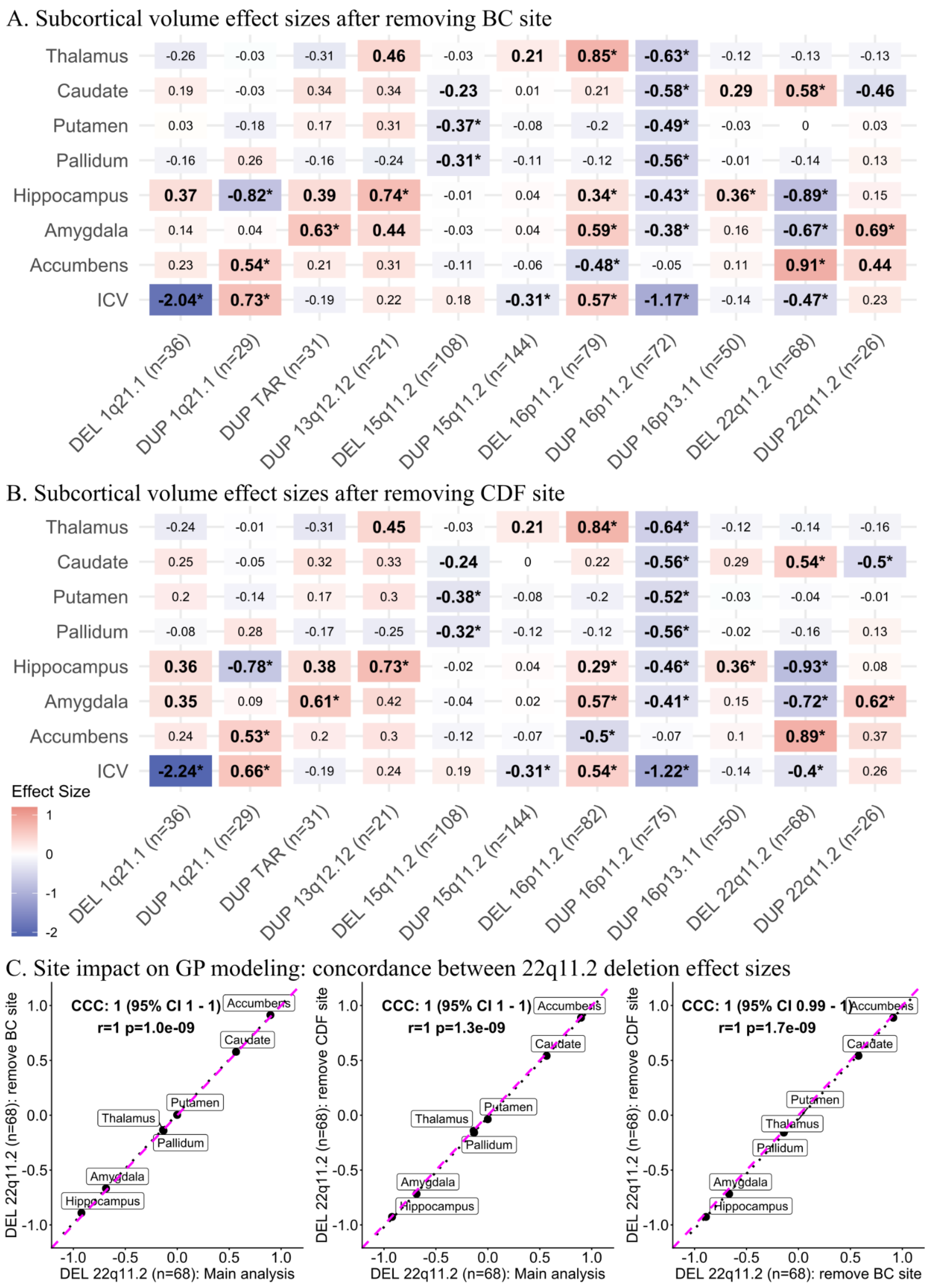


Supplement Figure 4: Impact of the site on Gaussian Processes modeling (leave one site out analysis).

Legend: A-B) Cohen's *d* values for subcortical structures and ICV for 11 CNVs. Leave-one-site-out analysis (Gaussian processes modeling and Cohen's *d* estimation) run after removing data from A) BC (Brain-Canada site); and B) CDF (Cardiff University site). Case-control differences were calculated (lm function in R) using W-scores (derived from Gaussian processes modeling). W-score already includes adjustments for age, sex, site, and ICV. Significant effect sizes with nominal p-value <0.05 are in bold, and FDR p-value <0.05 are shown with an asterisk (*); FDR correction was applied across all CNVs and structures. Darker red or blue represent higher positive or negative effect sizes. Sample sizes for each analysis (for ICV) are reported in parentheses along with x-axis labels. DEL: deletions; DUP: duplications; ICV: Intracranial volume. The main analysis results are reported in **Figure 1B**.

C) Concordance plots between 22q11.2 deletion effect sizes (Cohen's d) for our main analysis; removing BC, and CDF. The perfect concordance line is shown in magenta. Linear fitted lines are shown with black dots. r: Pearson correlation; p: p-value obtained using a parametric test (cor.test function in R); CI: confidence interval; CCC: concordance correlation coefficient (using *DescTools* package in R).

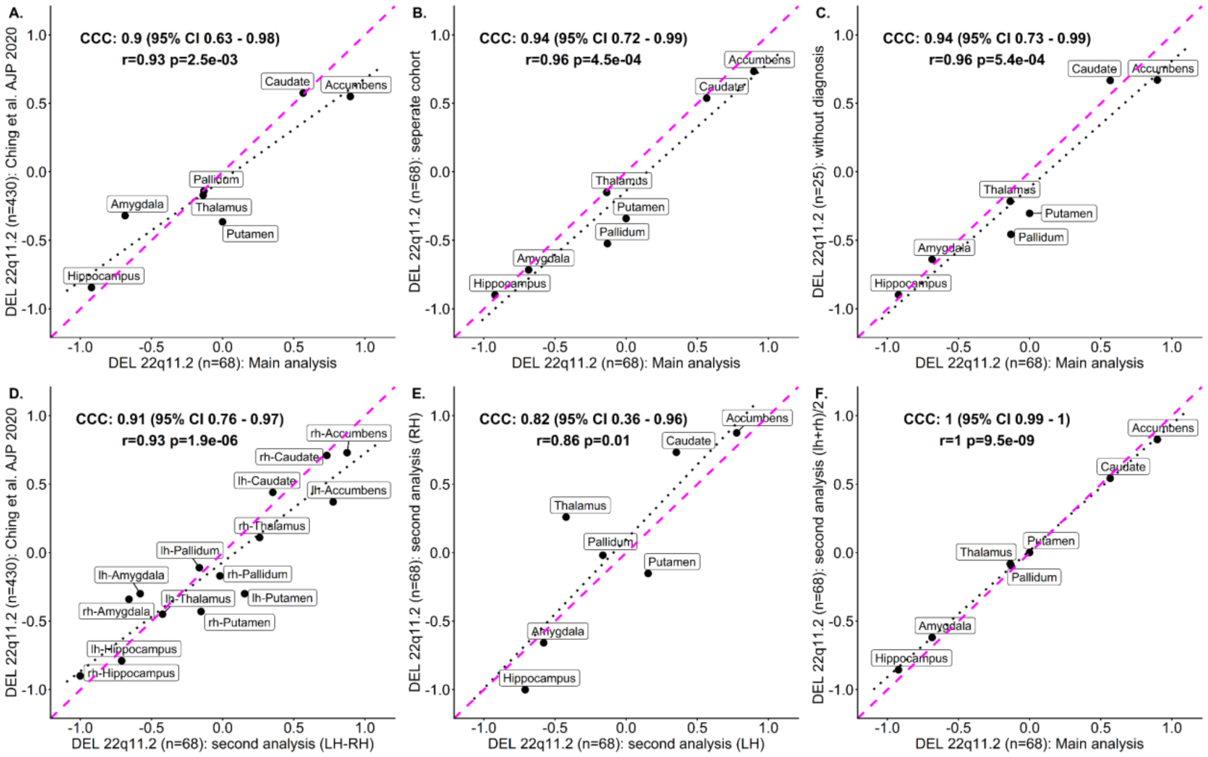


Supplement Figure 5: Sensitivity analysis subcortical volume effect sizes.

Legend: Concordance plots between 22q11.2 deletion effect sizes (Cohen's *d*) for our main analysis (7 ROIs) and different experiments. A) Concordance with literature: comparison with Cohen’s d values (average of left and right hemisphere) from Ching et al. 2020; B) Impact of data pooling: comparison with effect sizes estimated by running analysis case-control linear model in separate cohorts; C) Impact of diagnosis: comparison with effect sizes estimated after removing 22q11.2 deletion carriers with a diagnosis (linear regression within 22q11.2 data from UCLA); D) Impact of bilateral ROIs: comparison between bilateral effect sizes estimated in our analysis and those reported in Ching et a. 2020; E) Concordance between left and right hemisphere effect sizes in bilateral analysis; F) Concordance between effect sizes estimated in our main analysis (average left and right hemisphere volumes before modeling or adjusting for covariates) versus left and right hemisphere averaged effect sizes. The perfect concordance line is shown in magenta. Linear fitted lines are shown with black dots. r: Pearson correlation; p: p-value obtained using a parametric test (cor.test function in R); CI: confidence interval; CCC: concordance correlation coefficient (using DescTools package in R).

A. Surface Cohen's d maps

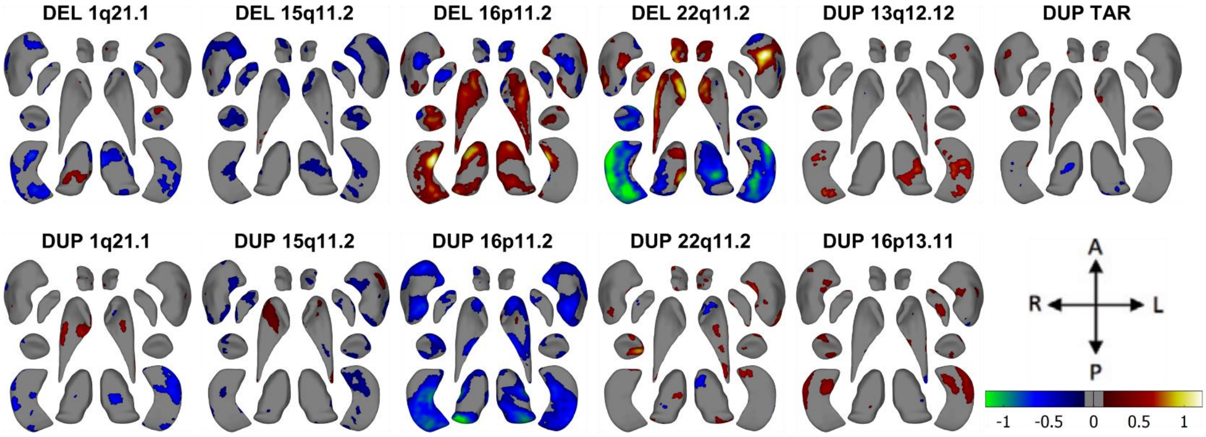


B. Thickness Cohen's d maps

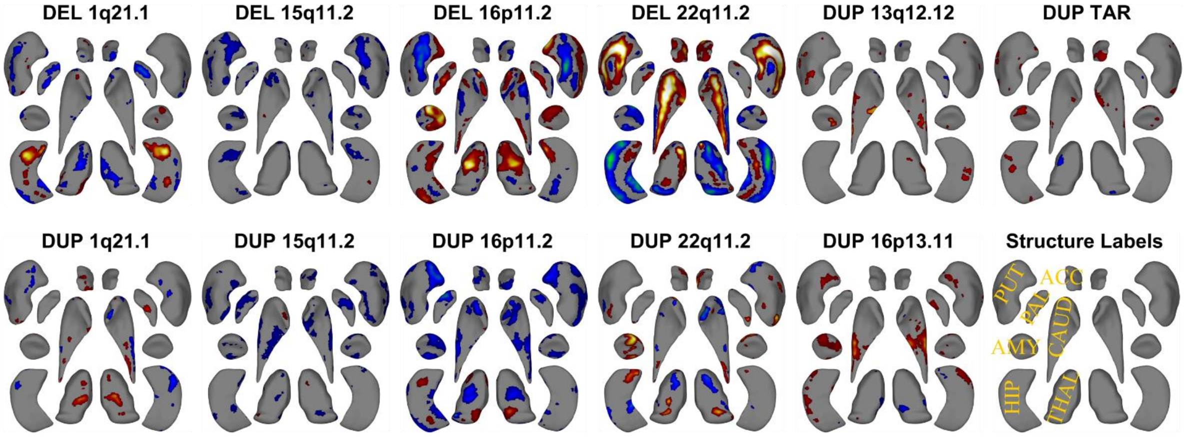


Supplement Figure 6: Cohen’s *d* maps for Subcortical Shape analysis (Ventral view)

Legend: A-B) Cohen’s d maps of subcortical shape alterations in surface (panel A); and thickness (panel B) for 11 CNVs (ventral view). Significant vertices, after applying FDR correction (<0.05) across all 27,000 vertices x 11 CNVs (within each panel), are shown. Colorbar for panels A-B are shown in panel A, and structure labels are shown in panel B. Thickness represents local radial distance, and surface represents local surface area dilation/contraction. Blue/green colors indicate negative coefficients or regions of lower thickness measures in the CNV group compared with the controls. Red/yellow colors indicate positive coefficients, or regions of greater thickness values in the CNV group compared with the controls. Gray regions indicate areas of no significant difference after correction for multiple comparisons. Each vertex was adjusted for sex, site, age, and intracranial volume (ICV). Dorsal views are shown in **Figure 2**.

Abbreviations, DEL: deletion; DUP: duplication; ACC: accumbens; AMY: amygdala; CAUD: caudate; HIP: hippocampus; PUT: putamen; PAL: pallidum; THAL: thalamus; Directions: L-left, R-right, A-anterior, P-posterior.

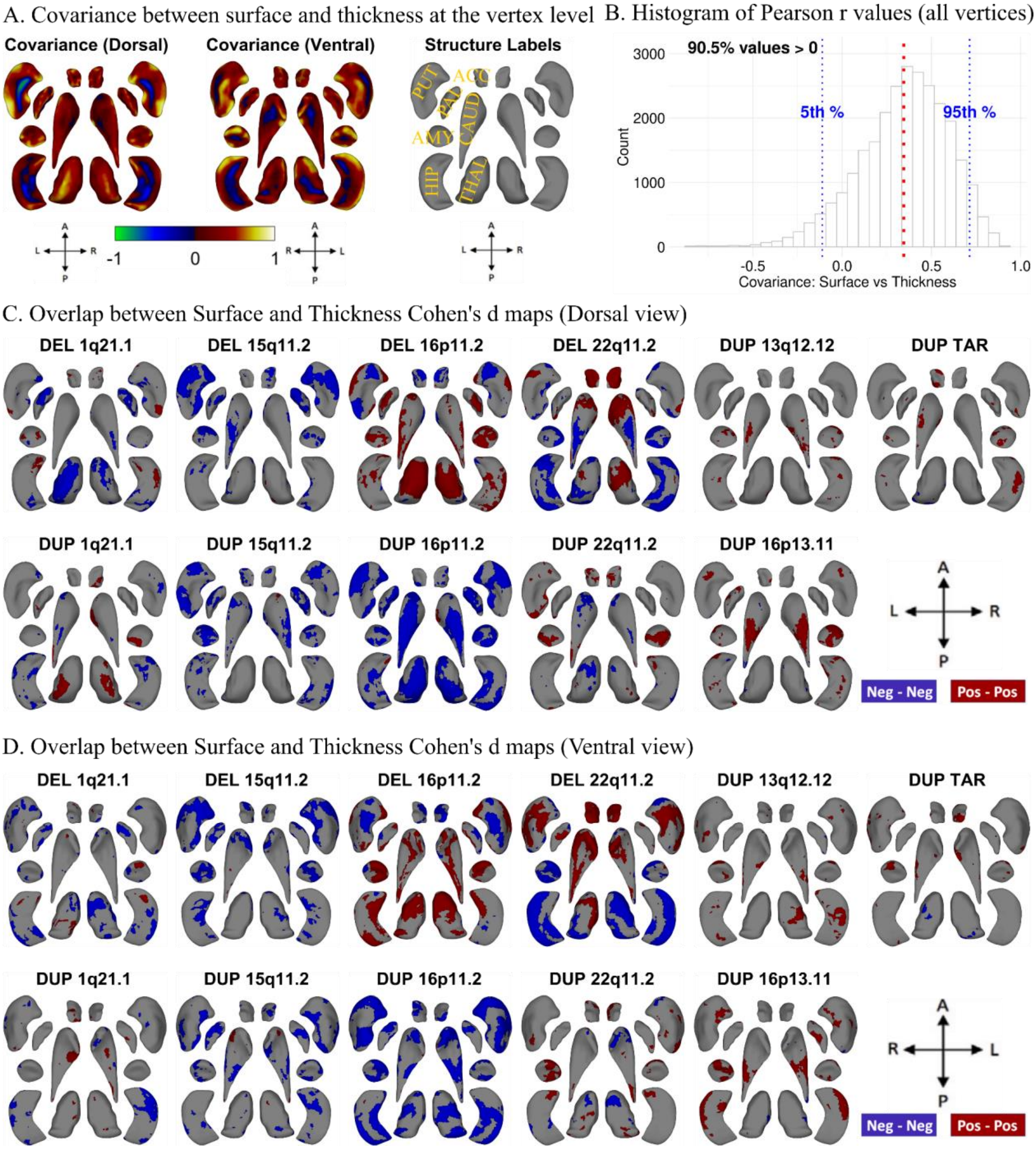


#### Supplement Figure 7: Covariance and overlap between surface and thickness at the vertex level

Legend: A-B) Maps of inter-individual variation between the local surface area and thickness. For each vertex, we computed a Pearson correlation between both metrics across all controls. The metrics were adjusted for age, sex, site, and ICV. A) The Pearson r values were projected on the dorsal and ventral maps of subcortical structures. B) The histogram of Pearson r values (X-axis) for each of the 27120 vertices across all subcortical structures demonstrates a strong bias towards a positive correlation between surface and thickness.

C-D) Overlap between surface and thickness vertices for each CNV; significant for either metric or both metrics: C) dorsal, and D) ventral views. Blue (Neg-Neg) and red (Pos-Pos) represent significant thinning and contraction or thickening and expansion respectively. For convex local curvature, overlapping/concordant alterations in the same vertices reflect subregional volume changes occurring near the structural surface. Neg and Pos refer to negative and positive Cohen's *d* values respectively.

Abbreviations, DEL: deletion; DUP: duplication; ACC: accumbens; AMY: amygdala; CAUD: caudate; HIP: hippocampus; PUT: putamen; PAL: pallidum; THAL: thalamus; Directions: L-left, R-right, A-anterior, P-posterior.

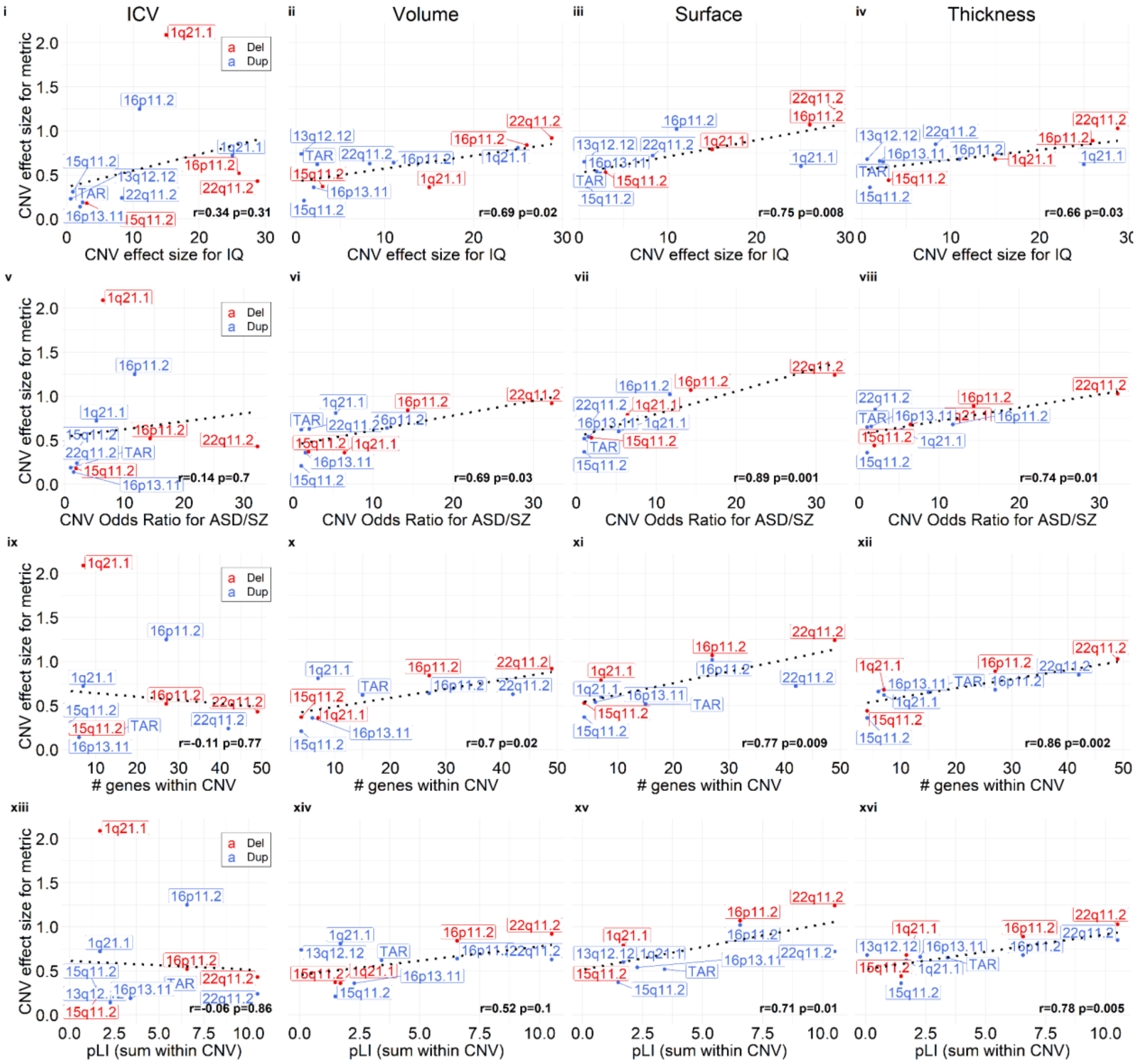


Supplement Figure 8: Comparison of effect sizes of CNVs on MRI-metric and disease-risk / cognition / #genes / pLI-sum.

Legend: Comparison of effect sizes of CNVs on ICV / subcortical-volume / subcortical-shape metrics (thickness and surface) and previously published effect sizes on cognition (first row), disease risk (second row), # genes within CNV (third row), and pLI-sum (fourth row). Regression lines fitted using the geom_smooth function in R. Pearson correlation and p-values (parametric) are reported within each plot. Y-axis: MRI-metric effect sizes (ES); X-axis: previously published effect-size on cognition; Odds Ratio for ASD/SZ; count of genes within CNV; and sum of pLI scores for genes within CNV. See **Supplement Table 2** for the effect sizes. Abbreviations, CNV: copy number variants; DEL: deletion; DUP: duplication;

NPD: neurodevelopmental and psychiatric disorders; Corr: Pearson correlation; ASD: autism spectrum disorder; SZ: schizophrenia; pLI: probability of being loss-of-function intolerant.

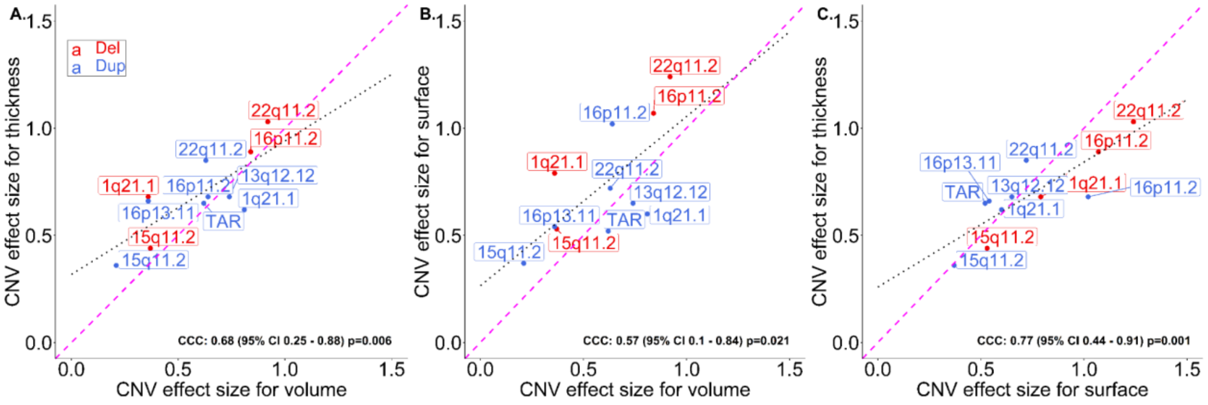


Supplement Figure 9: Concordance Correlation Coefficient between subcortical volume, thickness, and surface effect sizes.

Legend: Concordance plots between effect sizes for subcortical volumes, thickness, and surface. The perfect concordance line is shown in magenta. Linear fitted lines are shown with black dots. CNV labels are shown for deletion and duplication. Del: deletion; Dup: duplication; ES: effect size; CI: confidence interval; CCC: concordance correlation coefficient (using DescTools package in R).

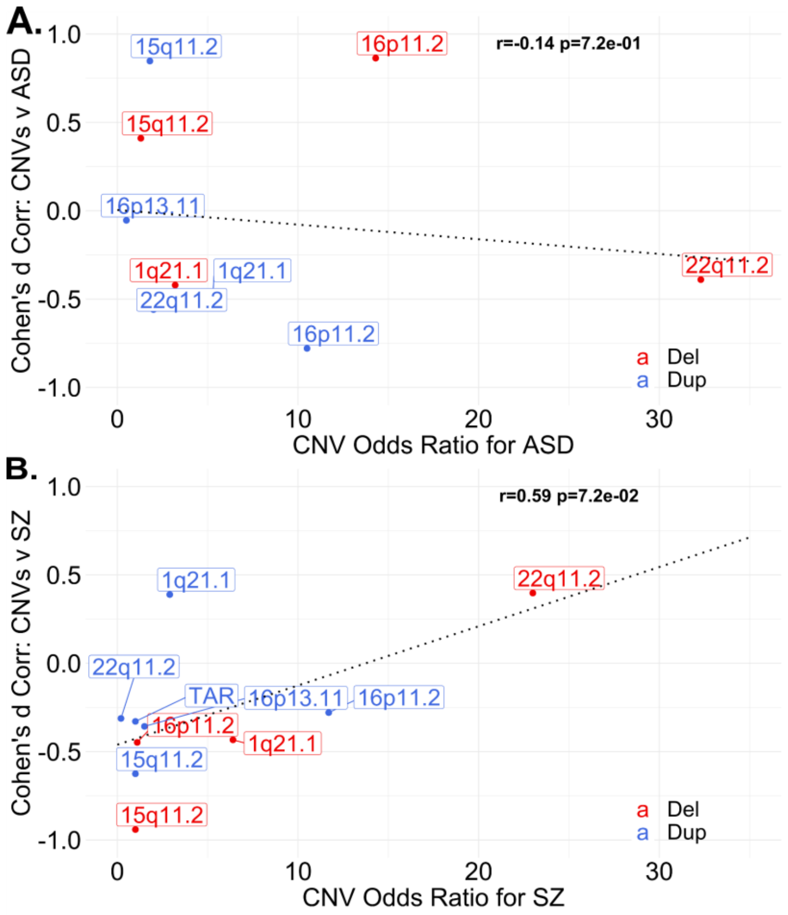


Supplement Figure 10: Correlation between Odds ratios for ASD/SZ, and Cohen's profiles' similarity of ASD/SZ with respective CNVs.

Legend: Pearson correlation and p-values between Odds ratios for ASD/SZ and subcortical volume alteration Cohen's profiles' similarity of ASD/SZ with the respective CNVs. X-axis: previously published Odds ratios for ASD and SZ (see **Supplement Table 2**); Y-axis: similarity in subcortical volume alteration Cohen's profiles': correlation between Cohen's *d* profiles of CNVs and ENIGMA's-Cohen's *d* profiles of ASD/SZ (see **Figure 3A**). Abbreviations, CNV: copy number variants; DEL: deletion; DUP: duplication; NPD: neurodevelopmental and psychiatric disorders; Corr: Pearson correlation; ASD: autism spectrum disorder; SZ: schizophrenia.

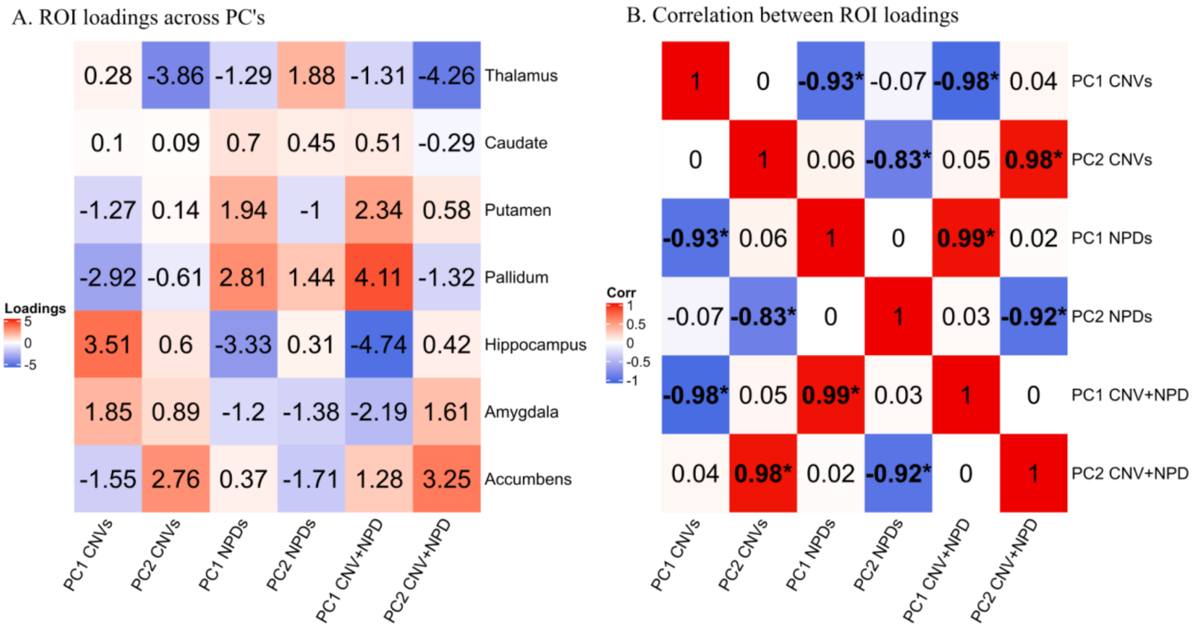


Supplement Figure 11: Latent dimensions across subcortical volumes of CNVs and NPDs.

Legend: Comparison of PCA ROI loadings across CNVs, NPDs, CNV+NPD. Panel A: ROI loadings across different PCA are shown. Panel B: Pearson Correlation between PCA ROI loadings are reported. PCA analyses were run using only 11 CNVs (n>20), only 6 NPDs: ASD, ADHD, BD, MDD, OCD, and SCZ; and 11 CNVs + 6 NPDs (as shown in **Figure 3**). BrainSMASH significant correlations are shown with *. Abbreviations, CNV: copy number variants; DEL: deletion; DUP: duplication; NPD: neurodevelopmental and psychiatric disorders; Corr: Pearson correlation; ASD: autism spectrum disorder; ADHD: attention deficit hyperactivity disorder; BD: bipolar disorder; MDD: major depressive disorder; OCD: obsessive-compulsive disorder; SZ: schizophrenia; PC: principal component; L: left hemisphere; Dim: dimension.

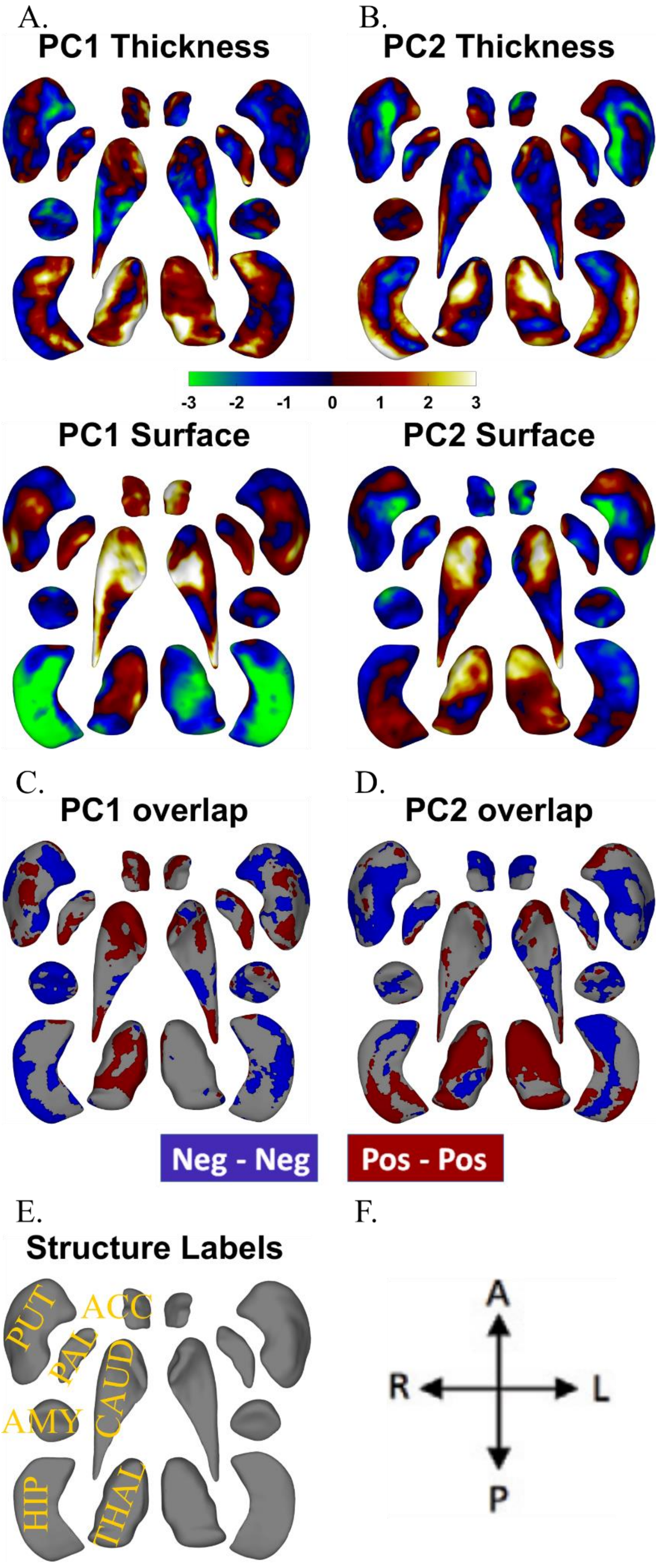
A.
PC1 Thickness
B.
PC2 Thickness
-3
-2
-1
0
1
2
3
PC1 Surface
PC2 Surface
C.
PC1 overlap
D.
PC2 overlap
Neg - Neg
Pos - Pos
E.
Structure Labels
PUT
ACC
PAL
CAUD
AMY
HIP
THAL
F.
A
R
L
P

#### Supplement Figure 12: Principal components analysis across vertex-wise Cohen's *d* maps of CNVs and NPDs (Ventral view).

Legend: Principal Component Analysis across vertex-wise Cohen's *d* maps of 11 CNVs and 2 NPDs (MDD and SZ) for thickness and local surface area. Thickness represents local radial distance, and surface represents local surface area dilation/contraction. Principal components analysis was run with CNVs as variables and vertices as observations (stacked across surface and thickness metric and all subcortical structures; Z-scored). For PC maps, blue/green and red/yellow colors indicate negative and positive coefficients respectively. For overlap maps, blue and red represent negative-negative / positive-positive thickness and surface PC loadings at each vertex respectively. Dorsal views are shown in **Figure 4**.

Abbreviations, DEL: deletion; DUP: duplication; PC: principal component; Dim: dimension; MDD: major depressive disorder; SZ: schizophrenia; ACC: accumbens; AMY: amygdala; CAUD: caudate; HIP: hippocampus; PUT: putamen; PAL: pallidum; THAL: thalamus; Directions: L-left, R-right, A-anterior, P-posterior.

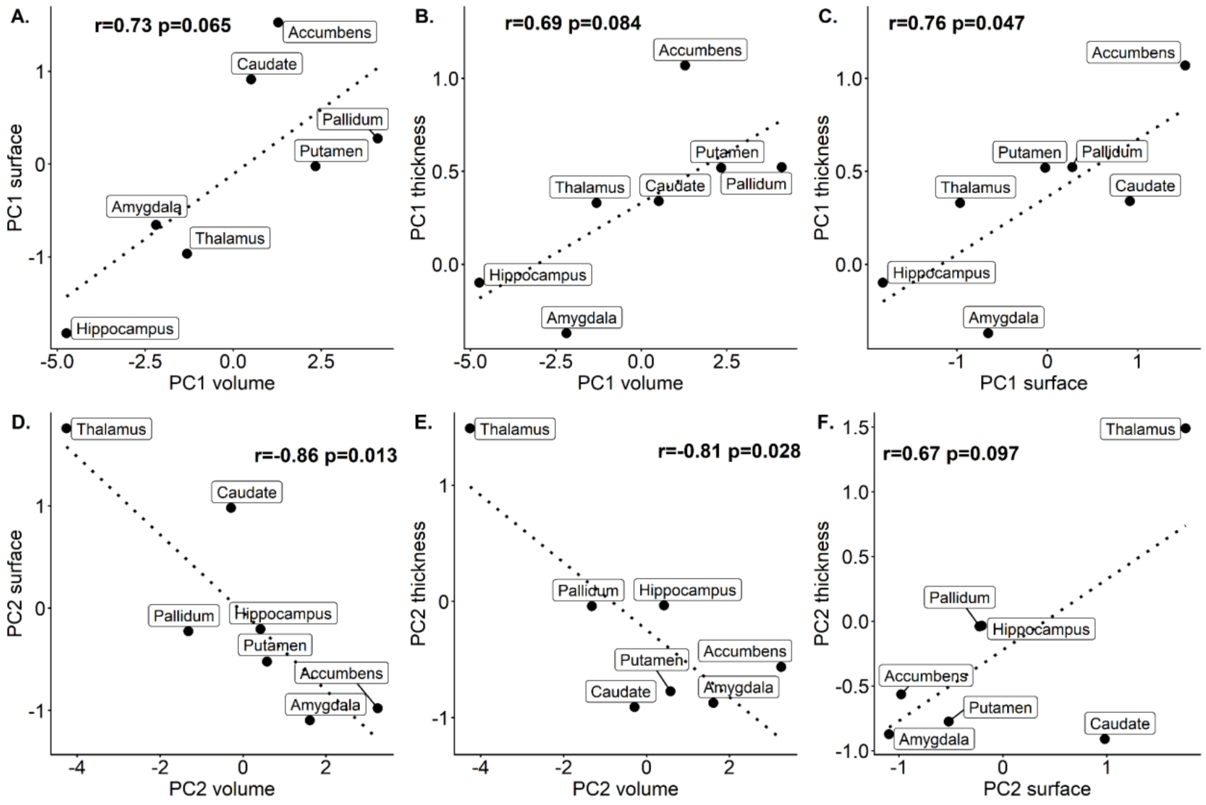


Supplement Figure 13: Correlation between latent dimension of subcortical structures identified across volume, thickness, and surface.

Legend: Correlation between PC1/PC2 loadings for subcortical volume (A-B, D-E) and PC1/PC2 loadings for thickness and local surface area. For subcortical shape metrics, thickness and surface, a summary value per structure is computed by taking the mean of respective PC loadings across all vertices in that structure.

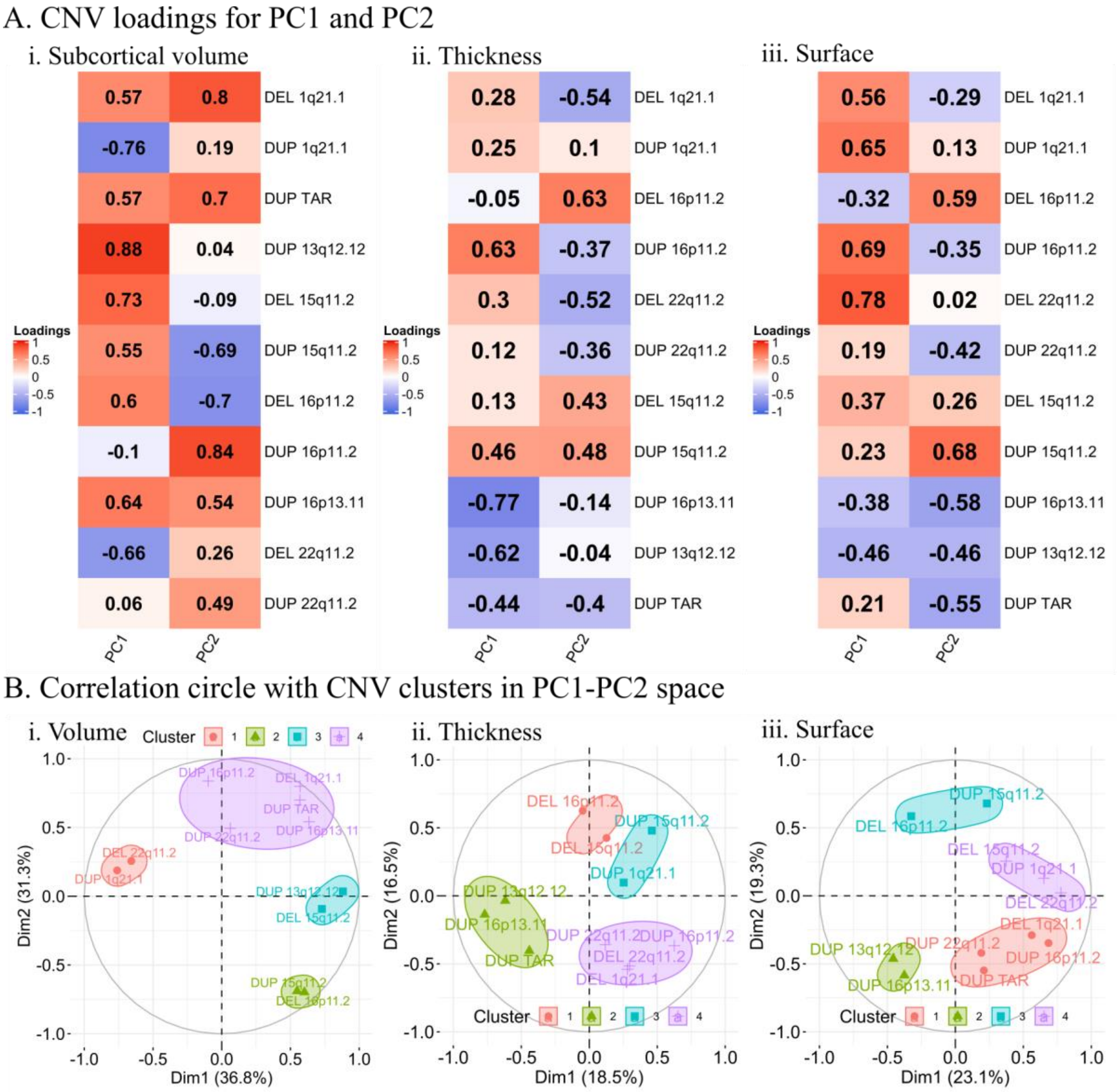


Supplement Figure 14: CNV only principal components analysis across the subcortical volume, thickness, and surface.

Legend: Panel A: CNV loadings for PCA analysis of Cohen's *d* maps of 11 CNVs for (i) volume (ii) thickness, and (iii) surface. Thickness represents local radial distance, and surface represents local surface area dilation/contraction. Principal Component Analysis was run with CNVs (n>20) as variables and volumes/vertices as observations (stacked across all subcortical structures and Z-scored).

Panel B: Correlation circle plot of CNV loadings in PC1-PC2 space with k-means clustering used to obtain k=4 groupings of CNVs. The percentage of variance explained by each principal component dimension is also included in axis labels.

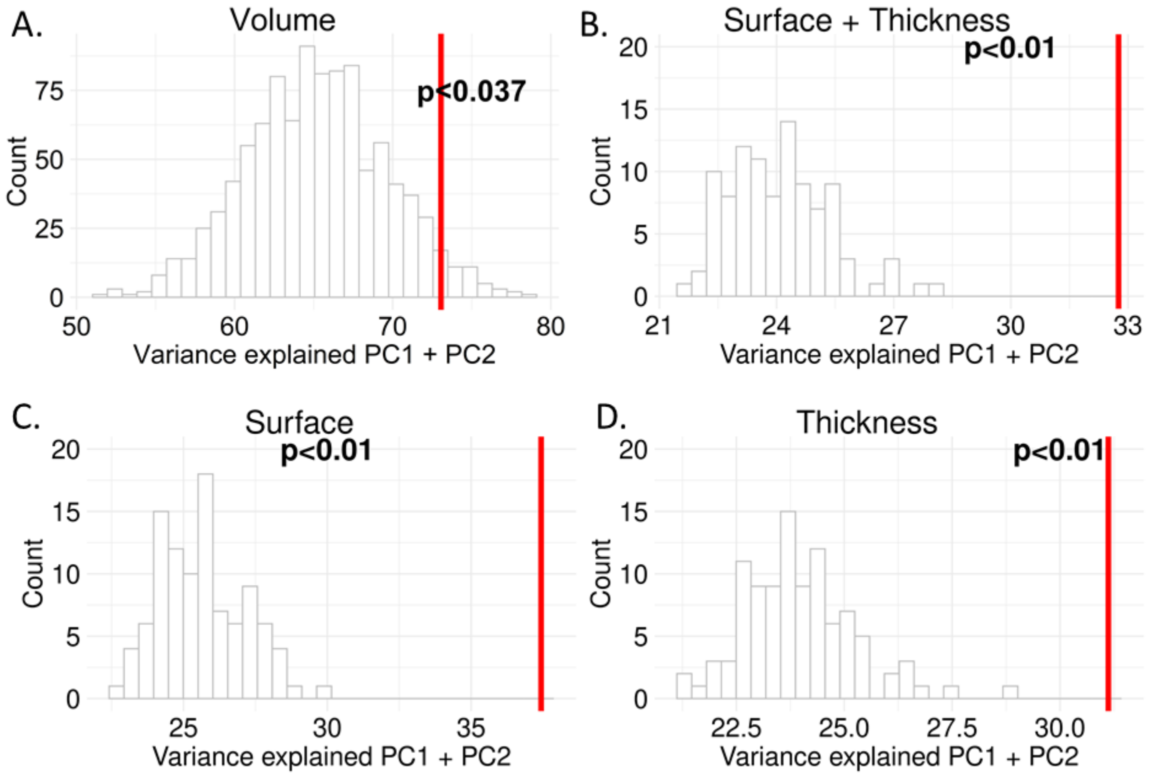


Supplement Figure 15: Statistical significance of variance explained by PC1 + PC2.

Legend: Variance explained by PC1 + PC2 w.r.t null distribution of variance explained (label shuffling) for A) subcortical volume; B) Surface and Thickness stacked; C) Surface; and D) Thickness. Volume null distribution was generated using 1000 label shuffles (CNV and control labels). For Surface + Thickness, Surface, and Thickness null distribution was generated using 100 label shuffles (CNV and control labels). X-axis: variance explained by PC1 + PC2; Y-axis: bin count.

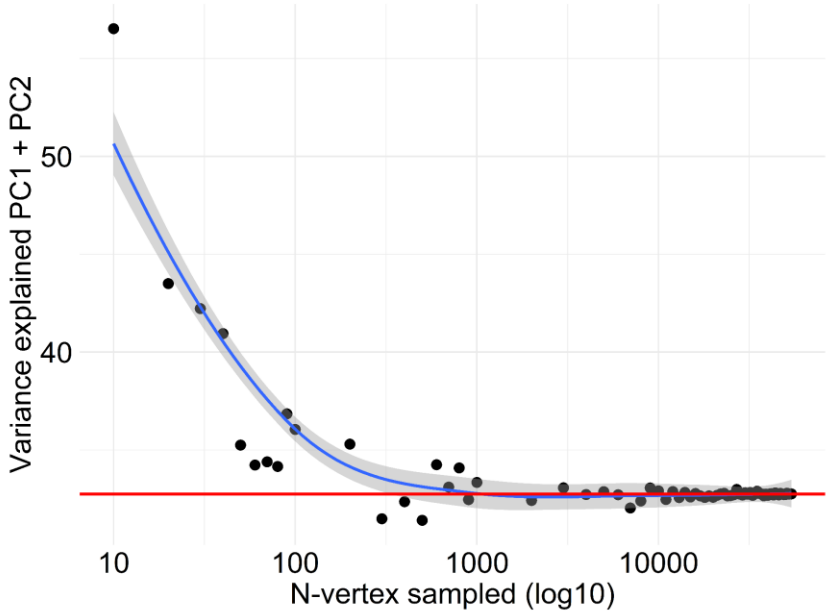


Supplement Figure 16: Variance explained in Shape analysis versus number-of-vertices.

Legend: Variance explained by PC1 and PC2 for surface + thickness (Y-axis) plotted against an increasing number of randomly sampled vertices (X-axis, log10 scale). The Red line shows the variance explained (32.8%) in the main analysis. The blue line and grey ribbon represent the *LOESS* fitting using geom_smooth function in R. N-vertex sampled ranges between 10 and 54000.
